# Multidomain Hypertension-Mediated Organ Damage in Ghanaian Adults: Prevalence, Correlates, and Brachial-Ankle Pulse Wave Velocity Performance

**DOI:** 10.64898/2026.05.28.26354393

**Authors:** K. O. Agyapong, E. Kyeremah, A. A. Folson, F. Agyekum, K. R. M. Blenman, L. T. Appiah, Y. Adu Boakye, I. K. Owusu

## Abstract

**Background:** Comprehensive assessment of hypertension-mediated organ damage (HMOD) across multiple organ systems in sub-Saharan Africa is limited. We assessed the prevalence and correlates of multidomain HMOD in a geographically diverse population in Ghanaian adult.

**Methods:** This cross-sectional secondary analysis of the Ghana Heart Study, which included 1,106 adults aged ≥18 years from four Ghanaian regions between September 2016 and March 2017. Multidomain HMOD was determined using a pre-specified 9-domain composite score ≥2, using an ESH/ESC 2018 guideline-informed selection of HMOD domain with baPWV instead of carotid-femoral PWV (cfPWV), due to device unavailability, and a threshold of ≥14 m/s which was derived from analysis within the cohort. LODO sensitivity analyses were used to address issues of predictor-outcome circularity. We used logistic regression models to examine association between each predictor and multidomain HMOD, adjusted for age, systolic blood pressure, body mass index, presence of dyslipidaemia and smoking status. We also performed receiver operating characteristic (ROC) analysis to determine correlates of multidomain HMOD and compare the discriminative ability of each predictor against the others.

**Results:** The mean age of participants was 46.9±17.2 years of which 58% were females. Multidomain HMOD was observed in 21.3% (235/1,106; zero-imputation lower bound 21.2%) of participants studied. There was a marked increase in the prevalence of multidomain HMOD with advancing age. Thus, while 8.6% (44/ 511) of adults<45years had multidomain HMOD, 20.6% (63/306) of 45- to 59-yr-olds and 44.4% (128/ 288) of individuals ≥60 years had multidomain HMOD. HMOD-positive adults were older (59.1±8.4 vs 43.6±13.4y, p<0.001), had higher systolic BP (147±22 vs 123±21 mmHg, p<0.001), and had higher prevalence of hypertension (73% vs 28%, p<0.001) than their HMOD-negative counterparts. Using the primary (circular) specification, the strongest co-occurrence among all domains of HMOD was observed between peripheral artery disease and other HMOD (OR 41.2, 95% CI 20.7-81.6; p<0.001) followed by valvular burden and other HMOD (OR 14.4, 95% CI 4.8-43.8; p<0.001) and between ECG-LVH and other HMOD (OR 9.0, 95% CI 5.9-13.8; p<0.001) (S2 Table). After LODO correction to remove the self-inclusive co-occurrence between each predictor domain and the outcome (all p-values calculated in S2 Table), there was no significant association between the remaining 8 HMOD domains and the prevalence of multidomain HMOD (all p-values>0.05; S2 Table). This was not the case for baPWV, however. Thus, whereas the AUC of the best performing non-self-inclusive HMOD domain (ECG-CMD) only reached 0.688±0.016 (vs 0.827±0.008 for self-inclusive AUC calculated for the sake of interest only and provided as supplementary material), baPWV demonstrated good discriminative capacity (LODO-adjusted AUC = 0.702, 95% CI 0.654-0.751; S3 Fig). However, this AUC did not significantly exceed that for age alone (AUC = 0.752; ΔAUC = −0.050; 95% CI −0.103 to 0.03; p=0.106; S3 Fig). Most importantly, after adjustment for SBP (a direct mediator in this pathway), the LODO AUC for baPWV did not exceed that for the single variable age (S3 Fig), indicating that baPWV does not possess independent discriminative power for multidomain HMOD above and beyond the information provided by SBP and age. Importantly, however, the adjusted OR for baPWV did not reach statistical significance (OR 1.094, 95% CI 0.986-1.213; p=0.091), suggesting that while circularity prevented validation of biological association, it did not prove the absence of association altogether. Sensitivity analysis (estimating total as opposed to direct effect) in which SBP was excluded from the regression model to estimate the total effect of baPWV on the prevalence of HMOD showed that, indeed, the OR for baPWV was significantly elevated (OR 1.261; 95% CI 1.150-1.382; p<0.001) in this specification. The effect of SBP, a direct mediator in this pathway, therefore apparently accounted for the non-significance in the original model entirely. Formal mediation analysis using the aforementioned specification yielded that SBP indeed mediated 69.9% (95% CI 41.3-128.8%) of the effect of baPWV on the prevalence of HMOD.

**Conclusions:** One in five Ghanaian adults has hypertension-mediated organ damage in multiple HMOD domains. baPWV has good discriminative power for HMOD risk prediction in a Ghanaian adult population under the non-circular LODO estimand (LODO-adjusted AUC = 0.702; 95% CI: 0.654, 0.751) than the PCE (AUC = 0.496; 95% CI: 0.438, 0.555; ΔAUC = +0.206; p < 0.001). However, baPWV LODO AUC (0.702) was not statistically significantly greater than age alone (AUC = 0.752; 95% CI: 0.730, 0.774; ΔAUC = −0.050; p = 0.106). AUC for self-inclusive model was provided in supplementary materials for the reader’s perusal, and that AUC (0.827; 95% CI: 0.794, 0.860) is circular. The prevalence of ECG-LVH was substantially higher (42%) than that of echocardiographic-LVH (5.9%) in this Black African population. These findings support further research on the role of baPWV for HMOD risk prediction in a Ghanaian adult population. Prospective validation of baPWV would be needed before clinical use.

## Introduction

Cardiovascular disease (CVD) is the leading cause of death globally with over 80% of mortality from CVD in low- and middle-income countries.^1,2^ The burden of CVD in Sub-Saharan Africa (SSA) is very high with western Sub-Saharan Africa having the highest age-standardized CVD prevalence of 9,475 per 100,000 population.^3^ Cardiovascular disease is increasing rapidly in many parts of Africa due to rapid epidemiological transition, urbanization, change in diet to more western type and increased sedentary behavior.^4,5^ In Ghana for instance, the rising prevalence of hypertension and related complications is a serious public health concern. The Ghana Heart Study (2016-2017) was a community-based cross-sectional study conducted in four regions of Ghana among 1,106 adults aged 18 years and above. The study found high age-standardized prevalence of 15.1% for obesity, 26.1% for hypertension, 6.8% for diabetes mellitus and 34.4% for dyslipidaemia.^6^ In terms of behavioral risk factors for CVD, 83.7% of the study participants were physically inactive, 81.4% had inadequate intake of fruits and 92% had two or more unhealthy behaviors.^7^ Hypertension can cause hypertension-mediated organ damage (HMOD) which are subclinical structural and functional changes in the heart, blood vessels, kidneys and other organs due to sustained high blood pressure.^8^ The detection of HMOD at an early stage is critical as it is a preclinical stage of cardiovascular disease and allows for time for intervention to prevent end organ damage. Previous work from the Ghana Heart Study found single organ HMOD or Total Organ Damage (TOD) prevalence of 10.1% for Peripheral Artery Disease (PAD), 8.3% for Carotid Artery Thickening, 4.1% for Left Ventricular Hypertrophy and 2.5% for Chronic Kidney Disease in a community based sample of Ghanaian adults.^6^ Importantly, hypertension frequently results in damage to more than one organ system and therefore multidomain HMOD or damage in two or more organ systems has not been previously characterized in SSA communities. However, most atherosclerotic cardiovascular disease (ASCVD) risk calculators developed for and validated in white populations have been found to perform poorly in Ghanaian and other West African populations. The degree of agreement among four commonly used ASCVD risk calculators, the Pooled Cohort Equations, Framingham Risk Score, WHO/ISH charts and Globorisk has been found to be poor to moderate (kappa = 0.3-0.8) ^9^. For instance, the use of the WHO/ISH charts, which are recommended by the CVD guidelines for Ghana for estimating cardiovascular risk ^10^, would classify about 82% of adults aged 40-79 years as of ‘low risk’ while the Framingham Risk Score would classify about 22% as of ‘high risk’ for cardiovascular events thereby necessitating intense treatment of hypertension and early intensification of treatment in contrast to the WHO/ISH charts. To our knowledge this is one of the first studies to apply a comprehensive ESH/ESC 2018 framework for the assessment of multiorgan hypertension-mediated organ damage (HMOD) in a community based sub-Saharan African (SSA) cohort. Prior SSA studies have examined single-organ damage (e.g., RODAM,^22^ CARRS^23^), but none have applied a multidomain ESH/ESC 2018-informed composite with circularity correction.

In this secondary analysis of the Ghana Heart Study, we aimed to: (1) determine the prevalence of multidomain HMOD in Ghanaian adults; (2) identify the strongest domain co-occurrences (recognising that domain-component correlates reflect composite construction rather than independent biological prediction) within the HMOD composite; (3) compare the discriminative performance of individual markers, particularly baPWV, against the ASCVD Pooled Cohort Equations for detecting multidomain HMOD; and (4) evaluate the incremental value of arterial stiffness measurement in hypertension risk stratification. These findings are interpreted together with previously published data on lifestyle risk factors and risk calculator performance from the same cohort, with the goal of informing targeted prevention strategies and the development of locally derived hypertension management models.^7,9^

## Methods

### Study Design and Population

This is a secondary analysis from the Ghana Heart Study, a community-based cross-sectional survey conducted between September 2016 and March 2017 among 1,106 adults (≥18 years) from eight communities (4 urban and 4 rural) in four of Ghana’s geographically, culturally and socioeconomically diverse regions (Greater Accra, Ashanti, Central and Northern regions). The study’s methodology has been described in detail previously.^6,7^ Adults (≥18 years) of Ghanaian citizenship who were resident in the study communities at the time of survey were included.

Excluded from the analysis were: pregnant women; subjects with Type 1 diabetes mellitus; and those with established atherosclerotic cardiovascular disease (suffered stroke, myocardial infarction or peripheral artery disease and reported as such in the study questionnaire); subjects with congenital heart disease; subjects with secondary cause of hypertension; and subjects who refused to give informed consent to participate in the study. Participants with the following conditions were excluded from the study: pregnancy, type 1 diabetes, established atherosclerotic cardiovascular disease (self reported stroke, myocardial infarction or peripheral artery disease), congenital heart disease, secondary hypertension and any other condition that was deemed to be outside the scope of the study. Participants who refused to give informed consent after full explanation of the study were also excluded. The study was registered at ChiCTR1800017374 and received ethical approval from the Committee on Human Research, Publications and Ethics of the Kwame Nkrumah University of Science and Technology and Komfo Anokye Teaching Hospital (CHRPE/AP/415/16). Written informed consent was obtained from all participants.

### Data Collection

The data for this study was gathered during the initial Ghana Heart Study, using set methods that were carried out by trained researchers, including medical officers and nurses, as explained in earlier reports.^6,7^ Demographic and lifestyle variables: Age, sex, ethnicity, education, marital status, employment status, region, and urban/rural residence were recorded. Smoking was defined as current use of any tobacco product (daily or occasionally). Physical inactivity was defined as <150 minutes of moderate-to-vigorous physical activity per week. Inadequate fruit and vegetable intake was defined as <2 servings per day, following WHO STEPwise approach criteria.^7,11^

Anthropometric and blood pressure measurements: To get accurate readings, people taking part in the study had their weight, height, and waist circumference measured while wearing light clothes and no shoes. Their body mass index, or BMI, was calculated by dividing their weight in kilograms by their height in meters squared. Blood pressure was checked three times after resting for 10 minutes, using a special device called an OMRON M6. The average of the last two readings was used to figure out if they had high blood pressure. High blood pressure, also called hypertension, was defined as having a systolic blood pressure of 140 mmHg or higher, or a diastolic blood pressure of 90 mmHg or higher, or currently taking medicine to lower blood pressure.^6^

Laboratory measurements: Fasting venous blood samples (8-12 hours overnight fast) were analyzed for total cholesterol, LDL cholesterol, HDL cholesterol, triglycerides, HbA1c, uric acid, hs-CRP, and creatinine. Dyslipidaemia was defined as TG ≥1.70 mmol/L, TC ≥6.22 mmol/L, LDL ≥4.14 mmol/L, HDL <1.04 mmol/L, use of lipid-lowering medications, or lipoprotein(a) ≥60 mg/dL.^6^

### Vascular Measurements

Arterial stiffness (baPWV): Brachial-ankle pulse wave velocity was measured using an OMRON/Colin VP-2000 automated oscillometric device.^21^ Measurements were taken from both limbs, and the mean of left and right values was used (or the single available value if only one side was valid). baPWV values are reported in meters per second (m/s). High baPWV was defined as ≥14 m/s. This threshold was derived from the study cohort as follows: the mean baPWV in the non-HMOD group was 11.4 m/s (SD 2.2), placing mean + 1 SD at 13.6 m/s, rounded to 14 m/s. The HMOD group mean was 14.9 m/s. This threshold is data-adaptive and derived from the study sample, which introduces optimisation bias into the self-inclusive AUC (0.827) but does not affect the LODO-adjusted AUC (0.702) because Domain 1 is excluded from the outcome in that analysis. No external West African normative baPWV data were available for validation (see Limitations).^21^

Peripheral artery disease (PAD) was defined as ankle-brachial index (ABI) <0.9 in either leg (strict inequality) using the same VP-2000 device that was used for the measurements of the ABI. All detected cases of PAD are cases of subclinical peripheral atherosclerosis since all participants with symptomatic PAD have been excluded from the study at baseline. Intima-media thickness (IMT) of the carotid arteries was measured by ultrasound (GE VIVID Q, 9L-RS probe). All IMT values reported in this manuscript represent the true intima-media thickness (mean 0.80 mm overall), not the outer vessel diameter. Carotid intima media thickness (IMT) was calculated for all far wall segments after the removal of sentinel values (-1) and outliers (>2.0 mm). Carotid plaques were defined according to the Mannheim Consensus criteria as focal intima media thickening of >1.5 mm or >0.5 mm above surrounding wall.^6^

### Echocardiographic Measurements

Echocardiographic Measurements Echocardiography: Transthoracic echocardiography was performed using a GE VIVID Q and 4S-RS probe by a trained cardiologist. Left ventricular mass index (LVMI) was calculated by using the Devereux formula with adjustment for body surface area.13 LVH on echocardiography was defined as LVMI > 95 g/m² in women and >115 g/m² in men.13 Left ventricular geometry was categorized as normal, eccentric or concentric remodeling or hypertrophy (concentric LVH or eccentric LVH). Concentric LVH was defined by increased LVMI with a RWT ≥0.42, whereas eccentric LVH was characterized by increased LVMI with a RWT <0.42. Diastolic function assessment (modified ASE/EACVI 2016 criteria): As above, assessment of diastolic function was conducted using a modified version of the ASE/EACVI 2016 criteria for assessment of diastolic function. In this work, assessment of left atrial enlargement used the left atrial linear dimension (>40mm) as a non-indexed parameter to approximate the threshold for increased left atrial volume index (LAVI>34mL/m²) as used in the 2016 ASE/EACVI guidelines. This may result in an overestimation of left atrial enlargement in individuals with central obesity. The fourth criterion for grading of diastolic dysfunction, peak TR velocity, was not included in this assessment as it results in an indeterminate classification in many individuals. Participants were therefore classified as having diastolic dysfunction or not on the basis of whether they met ≥2 of the following 3 criteria: 1) >septal and/or lateral e’ (tissue Doppler imaging) <7mm/s and 10mm/s respectively; 2) average E/E’ ratio >14; and 3) left atrial diameter >40mm (or LAVI >34mL/m²). Patients were then classified according to the 2016 ASE/EACVI recommendations for the grading of severity of diastolic dysfunction using echocardiography (Grade 1: impaired relaxation with E/A<0.8; Grade 2: pseudonormal with E/A 0.8-2.0; Grade 3: restrictive filling with E/A>2.0) and additional functional parameters, with the additional criterion of E/A ratio used for grading of severity of dysfunction rather than for classification into a specific category of diastolic dysfunction. The degree of valvular regurgitation was measured using a 0-4 score (0-none; 1-trace; 2-mild; 3-moderate; 4-severe) with scores of ≥3 classified as significant. 1. e’ septal <7 cm/s or e’ lateral <10 cm/s (TDI) (Figure 4). 2. Average E/E’ ratio >14 3. Left atrial diameter >40 mm (or left atrial volume index >34 mL/m²). Note: the E/A ratio is used for grading diastolic dysfunction into Grade I (impaired relaxation, E/A < 0.8), Grade II (pseudonormal, E/A 0.8-2.0) and Grade III (restrictive, E/A > 2.0) but not as a fourth criterion for the presence of diastolic dysfunction. Valvular regurgitation was graded from 0 (none) to 4 (severe) and participants with a grade of 3 or more were defined as having significant regurgitation.

### ECG Measurements

ECG-LVH was defined by Sokolow-Lyon (SV1+RV5/V6>3.5 mm) or Cornell (RaVL+SV3>2.8 mm in men, >2.0 mm in women) criteria.

### Renal Measurements

Estimated glomerular filtration rate (eGFR) was calculated using the CKD-EPI equation. Participants with eGFR <60 mL/min/1.73m² were classified into CKD stages ≥3. In addition, the urine albumin-to-creatinine ratio (UACR) was determined and categorized into microalbuminuria (30–299 mg/g) and macroalbuminuria (≥300 mg/g).

### Definition of Multidomain Hypertension-Mediated Organ Damage (HMOD)

We constructed a 9-domain composite score of hypertension-mediated organ damage (HMOD) based on the ESH/ESC 2018 hypertension guidelines framework^27^ and established pathophysiological links to hypertension-mediated organ injury. In summary, we defined each of the 9 organ system domains as either present (1) or absent (0). 1. High baPWV (≥14 m/s) - vascular stiffness (Note: the 2018 ESC/ESH guidelines designate carotid-femoral PWV [cfPWV] as the endorsed arterial stiffness metric; baPWV was substituted due to device availability and captures a physiologically distinct composite central-plus-peripheral arterial segment.). 2. Peripheral artery disease (ABI <0.9) - peripheral atherosclerosis 3. Carotid plaque (defined as focal IMT >1.5 mm or >0.5 mm above surrounding wall per Mannheim Consensus) - carotid atherosclerosis 4. ECG-LVH (Sokolow-Lyon or Cornell criteria) - electrical remodelling 5. Echocardiographic LVH (LVMI >115 g/m² men / >95 g/m² women) - structural cardiac damage 6. Diastolic dysfunction (modified ASE/EACVI 2016 criteria) - functional cardiac impairment. 7. Valvular damage – significant valvular regurgitation. 8. CKD stage ≥3 (eGFR <60 mL/min/1.73m²) - renal impairment 9. Albuminuria (UACR ≥30 mg/g) - renal microvascular damage For the purpose of this cross-sectional study, a patient was considered to have Multidomain HMOD when he or she had a score of 2 or more for different HMOD domains (i.e. vascular, peripheral artery disease, cardiac and renal HMOD). A study has used score of 2 or more for different European hypertension organ damage to divide patients into with high and low risk of cardiovascular morbidity and mortality in Europe.^28^ The results for this composite are presented in Fig. 6 for a score of 2 or more and in Fig. 7 for a score of 3 or more (as suggested in the European hypertension guideline documents). The number of cases for a score of 3 or more (i.e. three different HMOD domains affected) and therefore at highest risk for cardiovascular events are shown in parentheses. The chosen number of 2 or more HMOD domains (i.e. from different organs) for the present definition of Multidomain HMOD is clinically meaningful as it includes the possibility that all organ damage may reside within a single organ. In other words, the first 3 domains are of cardiac origin (ECG-LVH, structural cardiac damage [Echocardiographic LVH], functional cardiac impairment [Diastolic dysfunction]) while the remaining domains are of non-cardiac origin (vascular damage (High baPWV), peripheral atherosclerosis (Peripheral artery disease), renal microvascular damage (Albuminuria)). The number of 2 or more HMOD domains was chosen a priori, a copy of the protocol (dated) or the pre-analysis plan are available upon request.

#### Justification of the Multidomain HMOD Composite

Subclinical manifestations of cardiovascular disease, such as left ventricular hypertrophy, increased arterial stiffness, renal microalbuminuria and others have been well validated as surrogate end points for future cardiovascular events in several populations.^8, 27^ In the context of hypertension, subclinical organ damage in multiple organs (HMOD) indicates more severe disease and the need for more aggressive preventive strategies. Thus, assessing damage across organs in patients with hypertension has been recommended by ESH/ESC 2018 guideline.^27^ A prospective study is required to validate this definition of multidomain HMOD against clinical end points in sub-Saharan African populations.

### ASCVD Risk Score Calculation

We calculated 10-year ASCVD risk using the Pooled Cohort Equation (PCE) for the 565 participants aged 40-79 years, the age range for which the PCE was validated for use in PCE-based estimates of future event risk (14). Note that the PCE was derived using data from Black and White populations. Thus, in contrast to use of the PCE for estimates of future cardiovascular event risk, here we use the PCE to assess prevalent cardiovascular damage.

### Statistical Analysis

All analyses were conducted using R version 4.3.3 (48). For the continuous variables, the data were presented as mean (SD) or median (IQR) and compared between the groups using the Wilcoxon rank-sum test or the Kruskal–Wallis test, as appropriate. For the categorical variables, the data were presented as numbers and percentages and compared between the groups using the Pearson’s chi-square test or the Fisher’s exact test, as appropriate.

#### Logistic regression

Logistic regression models, adjusted for age (as a non-linear continuous variable, modeled with restricted cubic splines (RCS) with 4 knots at percentiles 10, 33, 67, 90; Likelihood Ratio Test χ²=8.88, df=2, p-value=0.012, hence RCS preferred over linear or 3-knot models), systolic BP, BMI, presence of dyslipidaemia and smoking were used to identify correlates of multidomain HMOD. The odds ratios and 95% confidence intervals of the correlates of the individual domain-components are provided. Here, these odds ratios reflect the co-occurrence of the individual domain-components with other components within the same multidomain construct, rather than independent effects of the individual domain-components. Leave-one-domain-out (LODO) sensitivity analyses were performed to estimate the effects of the individual domain-components that are truly external to the multidomain construct (i.e. Domain 1 LDH, Domain 5 HbA1c, and Domain 9 hs-CRP).

#### ROC analysis

Area under the receiver operating characteristic curve (AUC) for individual subclinical markers to assess their ability to discriminate between cases and controls (i.e. to predict the presence of multidomain HMOD) were calculated and compared using DeLong’s test.^30^ AUC values for all candidate subclinical markers to predict multidomain HMOD (baPWV, LVMI, eGFR, E/E’, ABI, IMT, hs-CRP, RVSP, EF and the 10-year ASCVD risk score) were calculated. Since each of the 9 HMOD domains can be considered in isolation (the composite is a ‘construct’ and the individual components or ‘domains’ are ‘objects’), an alternative AUC was also calculated for each of the individual 9 subclinical markers to predict the presence of any single HMOD domain (i.e. a self-inclusive AUC, supplementary Table S4). The LODO-adjusted AUC is the pre-specified primary estimate of a subclinical marker’s ability to predict multidomain HMOD status, while the self-inclusive AUC for all 9 subclinical HMOD domain-components provides supplementary information on their individual ability to predict each of the individual HMOD domains, in isolation.

#### Incremental value

None of the above calculations are appropriate for assessing the incremental value of any of the above predictors (or their continuous forms) in addition to the PCE estimate of 10-year ASCVD risk because the above-threshold dichotomized form of any predictor is a component of the multidomain HMOD construct defined above and thus the predictor would be added to a very poor baseline model (the PCE does not estimate prevalent damage, only future risk). Therefore, the above AUCs would be expected to increase substantially (on a theoretical basis, perhaps even by as much as or greater than AUC.900) and the resulting ΔAUC would not have any well established clinical interpretation. Supplementary Table S3 presents results from a poor but supplementary calculation to provide some insight.

#### Missing data

This was quantified for all variables and was found to be low for all variables except UACR where 36.8% of values were missing. All other variables had missing values for less than 10%. Thus complete-case analysis was used. For Domain 9, UACR, cases in which this variable was missing were assigned a score of zero (i.e. the case was assigned to the group without this component of HMOD). Little’s MCAR test (multivariate global test) was also used to test for missingness at random. Outcome-specific missingness was also examined for. As a sensitivity analysis, multiple imputation by chained equations (MICE) was also used to handle missing values. This used a number of auxiliary variables (age, systolic BP, BMI, diabetic status, eGFR) and created 20 complete data sets (m=20). All relevant data were available except for 36.8% of the UACR measurements. Little’s MCAR test (a multivariate global test) was used to assess whether the missing data were Missing Completely At Random (MCAR). In addition, the outcome-specific missingness was evaluated. To address the issue of missing data in a more robust manner, multiple imputation by chained equations (MICE) was performed using the five auxiliary variables age, systolic BP, BMI, diabetes status and eGFR (m = 20 imputations).

#### Model specification and diagnostics

The maximum number of parameters in the models used for analysis was 16. The events per variable (EPV) for the HMOD-positive cases was calculated by dividing the number of HMOD-positive events by the number of parameters in the respective model. The results for log(UACR) are displayed for the subgroup of participants with valid measurements of UACR in supplementary Table S3.

#### Circularity correction (LODO analysis)

SBP is a proximate mediator on the baPWV→HMOD pathway. Thus, we are interested in the direct effect of baPWV on HMOD. This effect is likely to be conservative and reduced as SBP is in the same pathway and likely to be highly correlated with the effect of interest. We performed a sensitivity analysis where we omitted SBP from the set of covariates that we adjusted for when estimating the effect of baPWV in the LODO analysis (see Table S2).

#### Interpretive note

Associations between case status and individual correlates within the primary model for correlates of multidomain HMOD are presented for informational purposes only, and it is essential to recognize that these reflect the co-occurrence of various domain components, and do not necessarily provide evidence of the independent relationship between a specific marker and HMOD. The three external domain components (LDL, HbA1c, hs-CRP) are all non-significant in the primary model. AUC values for self-inclusive models (i.e., all 9 domain components combined into a single threshold variable) are provided in supplementary materials for comparison purposes only and represent a variable’s ability to identify all cases of HMOD (all 9 organ damage categories combined into 1 threshold variable), and should not be used as an index of the construct’s validity as a multidomain composite in this study. LODO-adjusted AUC values provide non-circular discriminative information for individual correlates within the 9 domain components of multidomain HMOD studied here. A p-value <0.05 (two-sided) was considered statistically significant.

The corresponding author (K.O. Agyapong) had full access to all the data in the study and takes responsibility for its integrity and the data analysis

## Results

### Participant Characteristics

A total of 1,106 participants were included. The mean age was 46.9 (17.2) years, and 58.0% were female. The majority (80.4%) resided in urban areas, and 57.8% were of Akan ethnicity (Table 1).

**Table 1.**
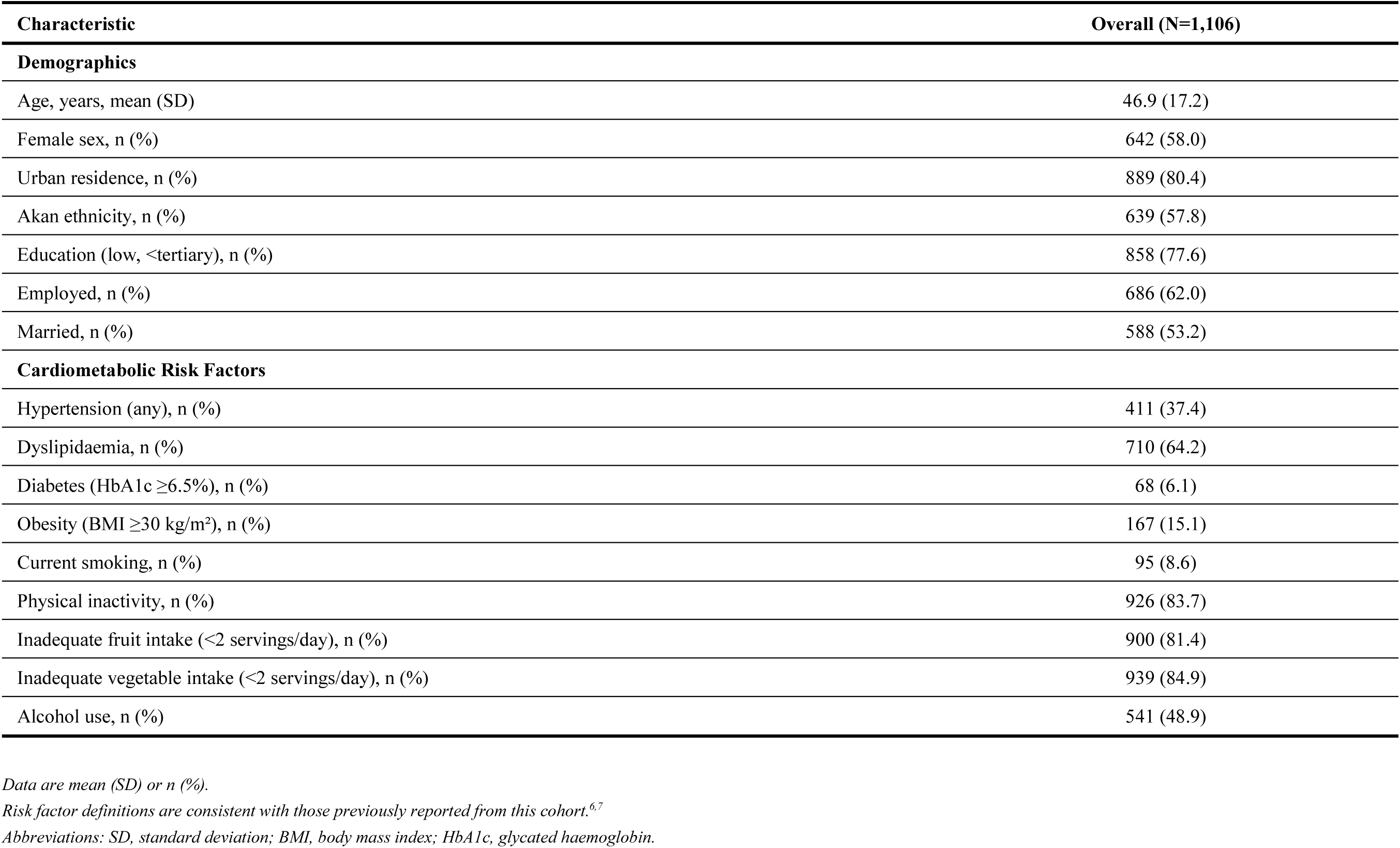
Baseline Characteristics of the Study Population (N=1,106)

| Characteristic | Overall (N=1,106) |
| --- | --- |
| **Demographics** |  |
| Age, years, mean (SD) | 46.9 (17.2) |
| Female sex, n (%) | 642 (58.0) |
| Urban residence, n (%) | 889 (80.4) |
| Akan ethnicity, n (%) | 639 (57.8) |
| Education (low, <tertiary), n (%) | 858 (77.6) |
| Employed, n (%) | 686 (62.0) |
| Married, n (%) | 588 (53.2) |
| **Cardiometabolic Risk Factors** |  |
| Hypertension (any), n (%) | 411 (37.4) |
| Dyslipidaemia, n (%) | 710 (64.2) |
| Diabetes (HbA1c $\geq 6.5\%$), n (%) | 68 (6.1) |
| Obesity (BMI $\geq 30$ kg/m<sup>2</sup>), n (%) | 167 (15.1) |
| Current smoking, n (%) | 95 (8.6) |
| Physical inactivity, n (%) | 926 (83.7) |
| Inadequate fruit intake (<2 servings/day), n (%) | 900 (81.4) |
| Inadequate vegetable intake (<2 servings/day), n (%) | 939 (84.9) |
| Alcohol use, n (%) | 541 (48.9) |

Data are mean (SD) or n (%).
Risk factor definitions are consistent with those previously reported from this cohort.<sup>6,7</sup>
Abbreviations: SD, standard deviation; BMI, body mass index; HbA1c, glycated haemoglobin.

### Prevalence of Multidomain Target Organ Damage

The composite HMOD score distribution is shown in Figure 1 and Table S1. Multidomain HMOD (composite score ≥2) was present in 235 participants (21.3%; zero-imputation lower bound 21.2%). Among those with multidomain HMOD, the majority had a score of 2 (66.0%) or 3 (24.7%), with 10.2% (24/235) having scores of 4 or 5 (representing 2.2% of the total cohort of 1,106).

**Figure 1.**
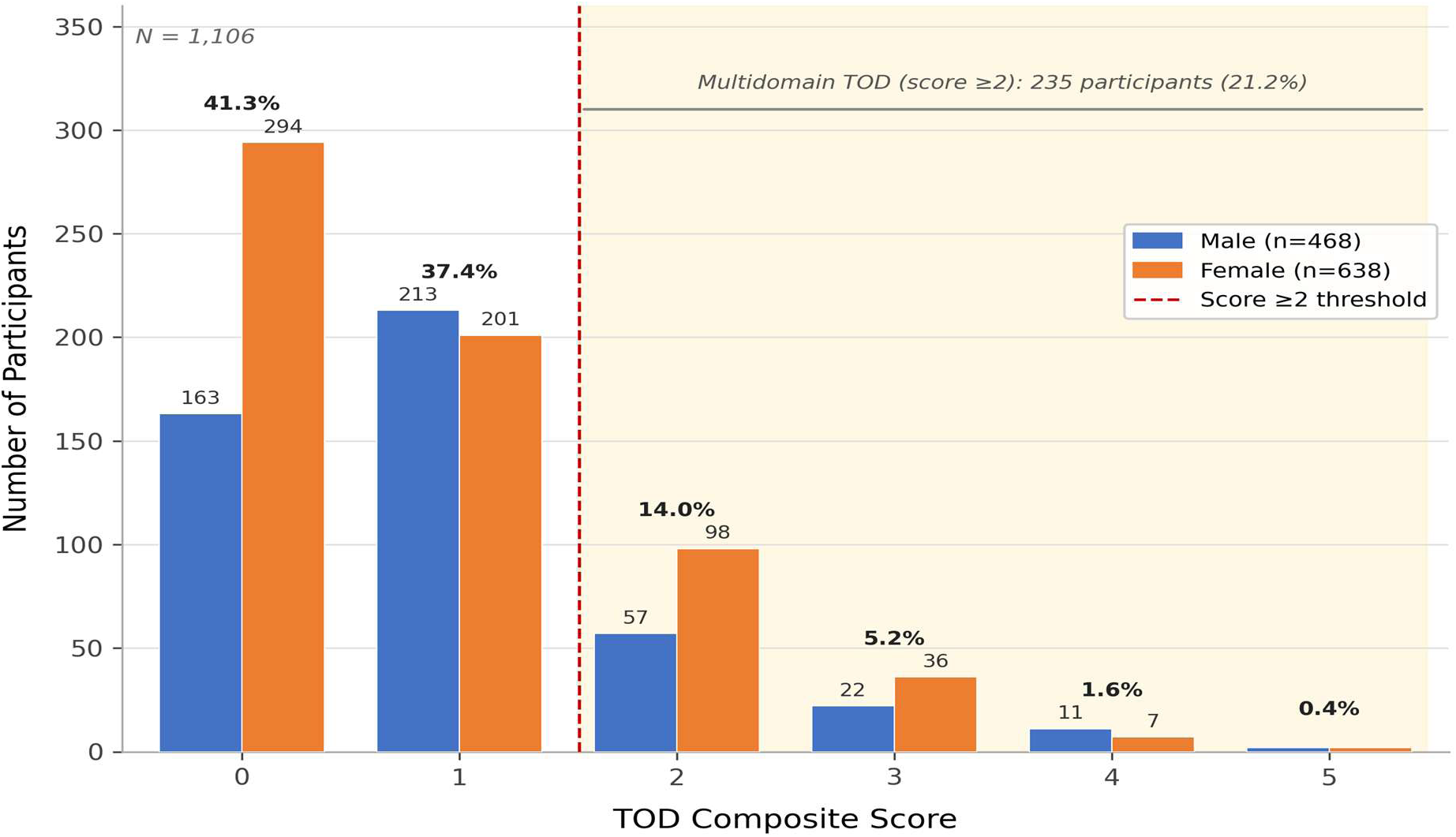
Distribution of the 9-Domain HMOD Composite Score (N=1,106). *Distribution of the 9-domain TOD composite score in 1,106 Ghanaian adults. Most participants had a score of 0 (41.3%) or 1 (37.4%), while 21.2% had multidomain HMOD (composite score ≥2). The distribution demonstrates that a substantial proportion of the population has evidence of subclinical damage across multiple organ systems.* *HMOD = hypertension-mediated organ damage (composite score). Multidomain HMOD defined as composite score ≥2 (TOD composite score)*.

### Characteristics by Multidomain HMOD Status

Participants with multidomain TOD were significantly older (59.1 (16.8) vs. 43.6 (15.8) years, p < 0.001) and had higher systolic blood pressure (147.1 (26.7) vs. 123.2 (18.9) mmHg, p < 0.001). Hypertension prevalence was nearly threefold higher in the TOD group (73% vs. 28%, p < 0.001). Dyslipidaemia was also more common (71% vs. 62%, p = 0.02). Complete characteristics across all domains are presented in Table 2.

**Table 2.** Participant Characteristics by Multidomain HMOD Status.

| Domain / Parameter | No HMOD (n=871) | HMOD (n=235) | p-value |
| --- | --- | --- | --- |
| **Demographics** |  |  |  |
| Age, years, mean (SD) | 43.6 (15.8) | 59.1 (16.8) | <0.001 |
| Male sex, n (%) | 374 (43) | 90 (38) | 0.2 |
| **Anthropometrics** |  |  |  |
| BMI, kg/m<sup>2</sup>, mean (SD) | 25.7 (5.8) | 26.4 (6.1) | 0.064 |
| Central obesity (waist >102/88 cm), n (%) | 278 (35) | 102 (44) | 0.006 |
| **Blood Pressure** |  |  |  |
| Systolic BP, mmHg, mean (SD) | 123.2 (18.9) | 147.1 (26.7) | <0.001 |
| Diastolic BP, mmHg, mean (SD) | 76.7 (11.5) | 86.9 (16.1) | <0.001 |
| Hypertension (any definition), n (%) | 240 (28) | 171 (73) | <0.001 |
| **Metabolic Risk Factors** |  |  |  |
| Dyslipidaemia, n (%) | 544 (62) | 166 (71) | 0.02 |
| Diabetes (HbA1c ≥6.5%), n (%) | 41 (4.8) | 27 (11.5) | 0.009 |
| **Vascular** |  |  |  |
| baPWV, m/s, mean (SD) | 11.4 (2.2) | 14.9 (3.0) | <0.001 |
| High baPWV (≥14 m/s), n (%) | 96 (11.0) | 139 (59.1) | <0.001 |
| ABI (minimum), mean (SD) | 1.02 (0.07) | 0.96 (0.09) | <0.001 |
| PAD (ABI <0.9), n (%) | 29 (3.3) | 66 (28.1) | <0.001 |
| Carotid IMT (true CIMT), mm, mean (SD) | 0.76 (0.23) | 0.95 (0.21) | <0.001 |
| Carotid plaque, n (%) | 2 (0.2) | 20 (8.5) | <0.001 |
| **ECG** |  |  |  |
| ECG-LVH (Sokolow-Lyon ≥3.5 mV), n (%) | 270 (31) | 166 (71) | <0.001 |
| ECG-LVH (Cornell >2.8/2.0 mV), n (%) | 44 (5.1) | 55 (23) | <0.001 |
| ECG-LVH (either criterion), n (%) | 284 (33) | 186 (79) | <0.001 |
| **Echocardiographic — Structure** |  |  |  |
| LVMI, g/m<sup>2</sup>, mean (SD) | 69.0 (15.2) | 83.9 (26.3) | <0.001 |
| Echo-LVH, n (%) | 7 (0.9) | 58 (25.0) | <0.001 |
| LV geometry, n (%) |  |  | <0.001 |
| — *Normal* | 524 (66) | 69 (30) |  |
| — *Concentric remodelling* | 261 (33) | 103 (45) |  |
| — *Eccentric LVH* | 1 (0.1) | 15 (6.5) |  |
| — *Concentric LVH* | 6 (0.8) | 43 (19) |  |
| LA enlargement (LAD >40 mm), n (%) | 119 (15) | 122 (53) | <0.001 |
| **Echocardiographic - Systolic Function** |  |  |  |
| Ejection fraction, %, mean (SD) | 65.9 (5.1) | 65.6 (6.0) | 0.8 |
| S' average TDI, cm/s, mean (SD) | 8.4 (1.6) | 7.8 (1.6) | <0.001 |
| Low S' (<7.5 cm/s), n (%) | 224 (26) | 100 (43) | <0.001 |
| **Echocardiographic - Diastolic Function** |  |  |  |
| E/A ratio, mean (SD) | 1.5 (5.0) | 0.9 (0.5) | <0.001 |
| Impaired relaxation (E/A <0.8), n (%) | 135 (17) | 133 (58) | <0.001 |
| E/E' average, mean (SD) | 7.4 (2.0) | 9.2 (3.4) | <0.001 |
| Elevated E/E' (>14), n (%) | 2 (0.2) | 19 (8.1) | <0.001 |
| Low E' (septal <7 or lateral <10), n (%) | 229 (26) | 173 (74) | <0.001 |
| Diastolic dysfunction (ASE 2016 criteria)<sup>†</sup>, n (%) | 2 (0.2) | 27 (11) | <0.001 |
| **Right Heart** |  |  |  |
| RVSP, mmHg, mean (SD) | 11.2 (0.5) | 11.1 (0.4) | <0.001 |
| Pulmonary hypertension (RVSP >36), n (%) | 1 (0.1) | 3 (1.3) | 0.03 |
| **Valvular** |  |  |  |
| Any significant regurgitation (≥moderate), n (%) | 6 (0.7) | 16 (6.8) | <0.001 |
| Valvular burden score, n (%) |  |  | <0.001 |
| — 0 | 865 (99) | 219 (93) |  |
| — 1 | 6 (0.7) | 13 (5.5) |  |
| — 2 | 0 (0) | 3 (1.3) |  |
| **Renal** |  |  |  |
| eGFR, mL/min/1.73m<sup>2</sup>, mean (SD) | 85.4 (7.0) | 77.5 (15.5) | <0.001 |
| CKD stage ≥3 (eGFR <60), n (%)<sup>*</sup> | 2 (0.3) | 34 (14.5) | <0.001 |
| UACR, mg/g, mean (SD) | 1.9 (4.9) | 12.2 (51.3) | <0.001 |
| Microalbuminuria (30–299 mg/g), n (%)<sup>*</sup> | 3 (0.4) | 12 (5.1) | <0.001 |
| **Composite Scores** |  |  |  |
| ASCVD 10-year risk, %, mean (SD) | 22.6 (27.7) | 23.3 (24.9) | 0.13 |
| High ASCVD risk (≥7.5%), n (%) | 387 (44) | 117 (50) | 0.2 |
| Composite Z-score, mean (SD) | −0.2 (0.5) | 0.6 (0.7) | <0.001 |

Data are mean (SD) or n (%). HMOD = hypertensive-mediated organ damage (composite score ≥2).
eGFR measured in 915 participants; CKD stage ≥3 present in 36 participants (3.3% of total cohort). UACR measured in 699 participants.
IMT measured in 1,058 participants (870 No HMOD, 188 HMOD). Valvular burden score = number of valves with grade ≥3 (moderate–severe) regurgitation (range 0–2).
<sup>†</sup> Echocardiographic diastolic/functional parameters available in n=792 No-HMOD and n=230 HMOD-positive (denominators for diastolic rows).
<sup>‡</sup> UACR measured in 570 No-HMOD (65%) and 129 HMOD-positive (55%); n=699 total; differential missingness rate p=0.004; UACR-derived percentages use measured denominators.
<sup>\*</sup> eGFR measured in n=915. Significant regurgitation defined as any valve graded ≥3, consistent with ESH/ESC 2018 guidelines<sup>27</sup>.
<sup>§</sup> RVSP values are reported as tricuspid regurgitation peak gradient (TRPG) only; right atrial pressure (RAP, typically 5–8 mmHg) was not added. Normal TRPG is <12 mmHg, consistent with these values. Standard echocardiographic RVSP = TRPG + RAP would be approximately 16–19 mmHg.

### Adjusted Odds Ratios for Multidomain Hypertension-Mediated Organ Damage

Multivariable logistic regression adjusting for age, systolic BP, BMI, dyslipidaemia, and smoking revealed the strongest independent associations with multidomain TOD (Figure 2; Table 3).

**Figure 2.**
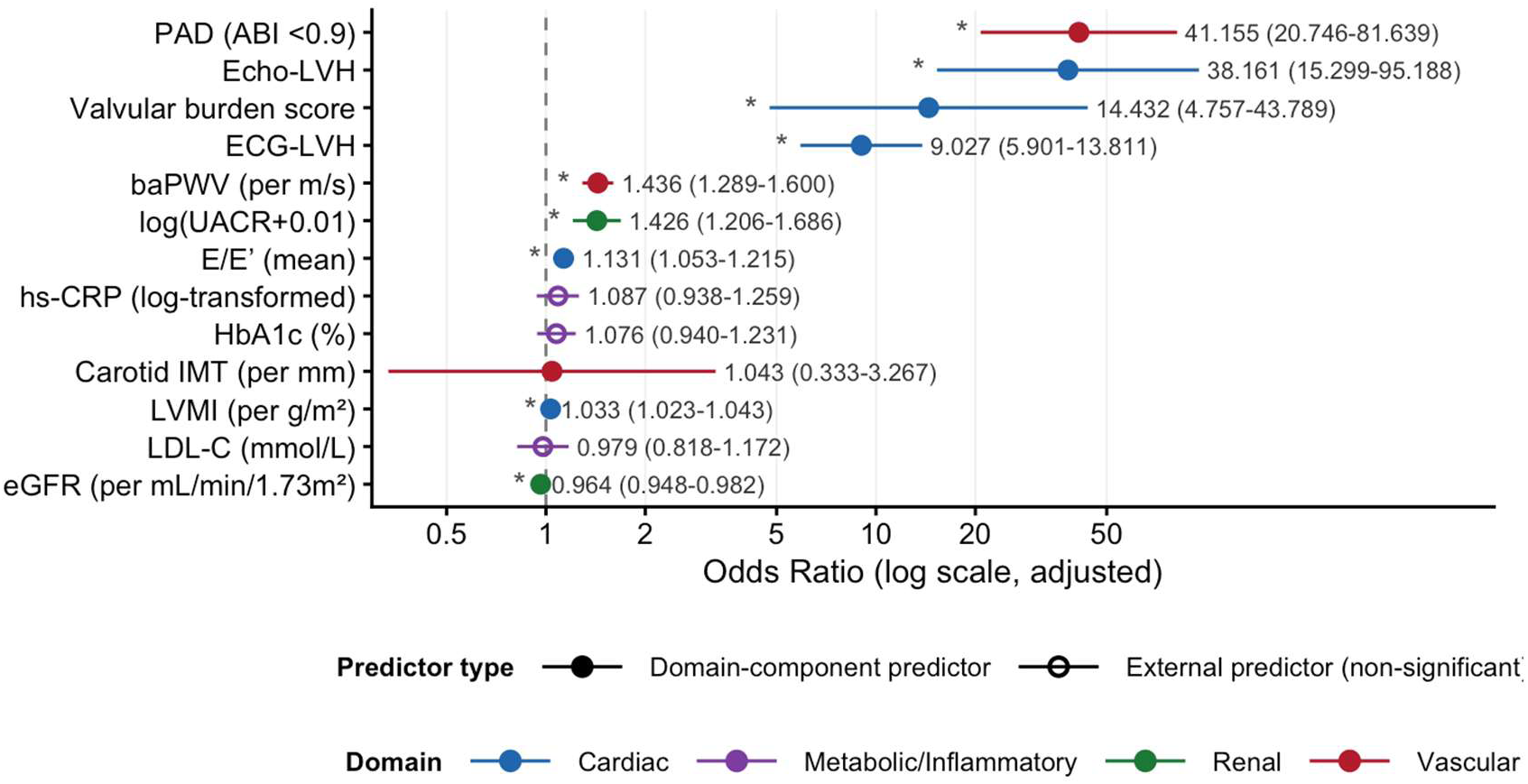
Adjusted Odds Ratios for Multidomain Hypertension-Mediated Organ Damage. *Forest plot showing multivariable-adjusted odds ratios (95% CI) for correlates of multidomain HMOD. Models were adjusted for age (restricted cubic splines, 4 knots), systolic blood pressure, BMI, dyslipidaemia, and smoking. ORs are displayed on a log scale and colour-coded by organ domain. PAD, valvular burden score, and ECG-LVH showed the strongest domain co-occurrences in the primary (circular) specification; LODO-corrected ORs for PAD (0.59, p=0.63) and ECG-LVH (1.52, p=0.09) were non-significant (see Table 6).* Only variables from the primary multivariable model (Table 3) are displayed. *ABI, ankle-brachial index; baPWV, brachial-ankle pulse wave velocity; CI, confidence interval; ECG-LVH, electrocardiographic left ventricular hypertrophy; eGFR, estimated glomerular filtration rate; E/E’, ratio of early mitral inflow velocity to early diastolic mitral annular tissue velocity; HbA1c, glycated haemoglobin; hs-CRP, high-sensitivity C-reactive protein; IMT, intima-media thickness; LDL-C, low-density lipoprotein cholesterol; LVMI, left ventricular mass index; OR, odds ratio; PAD, peripheral artery disease; UACR, urine albumin-to-creatinine ratio*.

**Table 3.** Multivariable-Adjusted Odds Ratios for Multidomain HMOD (TOD Composite Score ≥2) All models adjusted for age (restricted cubic splines, 4 knots; primary specification), systolic BP, BMI, dyslipidaemia, and smoking. N=1,106. Events=235. EPV=14.7 (235/16 parameters). Domain-component correlates (Sections A–C) reflect composite construction; LODO-corrected ORs (column 4) are the appropriate independent-association estimates.

| Predictor | OR (95% CI)<sup>a</sup> | p-value | LODO OR (95% CI)<sup>b</sup> | LODO p | Notes |
| --- | --- | --- | --- | --- | --- |
| **A. Vascular Domain Correlates** |  |  |  |  |  |
| baPWV (m/s, continuous)<sup>a</sup> | 1.44 (1.32–1.57) | <0.001 | 1.094 (0.986–1.213) | 0.091 | Self-inclusive AUC 0.827; LODO AUC 0.702 [primary] |
| ABI (PAD, <0.9)<sup>b</sup> | 41.2 (20.7–81.6) | <0.001 | 0.586 (0.065–5.271) | 0.634 | Domain co-occurrence; LODO non-significant |
| Carotid IMT (mm) | 45.6 (18.2–114.3) | <0.001 | 0.704 (0.663–0.745)<sup>†</sup> | — | <sup>†</sup>AUC, not OR (Domain 3=plaque, IMT non-circular) |
| **B. Cardiac Domain Correlates** |  |  |  |  |  |
| ECG-LVH (any criterion)<sup>b</sup> | 9.03 (5.87–13.88) | <0.001 | 1.519 (0.941–2.454) | 0.087 | Domain co-occurrence; LODO non-significant |
| *Sokolow-Lyon $\geq 3.5$ mV* | — | — | — | — | Component criterion (not in primary model) |
| *Cornell $> 2.8/2.0$ mV* | — | — | — | — | Component criterion (not in primary model) |
| Echo-LVH<sup>b</sup> | 38.16 (15.30–95.19) | <0.001 | — | — | Firth check: see Table S2; domain co-occurrence |
| E/E' average | 1.15 (1.08–1.23) | <0.001 | 0.656<sup>†</sup> | — | <sup>†</sup>LODO AUC |
| LVMI (g/m<sup>2</sup>) | 1.023 (1.014–1.032) | <0.001 | 0.616<sup>†</sup> | — | <sup>†</sup>LODO AUC |
| Valvular burden score<sup>c</sup> | 13.45 (4.76–42.77) | <0.001 | 0.515<sup>†</sup> | — | <sup>†</sup>LODO AUC; Firth penalized; MLE OR 14.43 (ratio 1.07) |
| **C. Renal Domain Correlates** |  |  |  |  |  |
| eGFR (mL/min/1.73m<sup>2</sup>) | 0.983 (0.976–0.990) | <0.001 | 0.618<sup>†</sup> | — | <sup>†</sup>LODO AUC |
| log(UACR+0.01)<sup>d</sup> | 1.43 (1.21–1.69) | <0.001 | — | — | n=699 subsample; EPV=8.1; see Table S5 |
| **D. External Correlates (Non-Domain Components)** |  |  |  |  |  |
| LDL cholesterol (mmol/L) | 1.05 (0.88–1.26) | 0.57 | — | — | Non-significant |
| HbA1c (%) | 1.06 (0.97–1.16) | 0.22 | — | — | Non-significant |
| hs-CRP (log, mg/L) | 1.08 (0.94–1.23) | 0.29 | — | — | Non-significant |

<sup>a</sup> Adjusted for age (RCS, 4 knots), SBP, BMI, dyslipidaemia, smoking. EPV = 235/16 = 14.7. Domain-component ORs reflect composite construction (co-occurrence effects), not independent biological prediction; LODO-corrected ORs are the appropriate independent-association estimates.
<sup>b</sup> Domain co-occurrence predictor; LODO OR is the non-circular estimate (Domain excluded from composite outcome).
<sup>c</sup> Firth penalized logistic regression applied (n=22 events). Near-identical Firth and MLE ORs reflect sparse data constraints, not methodological robustness; CI (4.76–42.77) indicates genuine instability.
<sup>d</sup> Firth penalized logistic regression for PAD (ABI <0.9, n=95 events): OR 39.18 (95% CI 20.26–78.78), p<0.001; MLE/Firth ratio 1.05. <sup>e</sup> Firth penalized logistic regression for Echo-LVH (n=65 events): OR 35.02 (95% CI 15.19–91.87), p<0.001; MLE/Firth ratio 1.09. <sup>f</sup> log(UACR+0.01) from n=699 subsample (168 HMOD-positive; EPV=168/16=10.5). Results in Table S5. OR may underestimate true association given differential UACR missingness (HMOD-positive: 55% measured vs 65% negative, p=0.004).

PAD (ABI<0.9) had the highest OR (41.2, 95% CI 20.7-81.6), followed by valvular burden (OR 14.4, 95% CI 4.8-43.8) and ECG-LVH (OR 9.0, 95% CI 5.9-13.8). baPWV was also significantly associated (OR 1.44 per m/s, 95% CI 1.32-1.57, p<0.001). In contrast, traditional metabolic markers (LDL, HbA1c, hs-CRP) were not significant after adjustment.

PAD (OR 41.2), valvular burden (OR 14.4), and ECG-LVH (OR 9.0) showed the strongest associations in the primary model. However, these correlates are domain components of the HMOD composite. LODO-corrected analyses showed substantial attenuation, with non-significant associations for PAD (LODO OR 0.59, p=0.63) and ECG-LVH (LODO OR 1.52, p=0.09), indicating that the primary ORs largely reflect domain co-occurrence within the composite rather than independent biological prediction.

### Discriminative Performance of Individual Markers

ROC analysis for the non-composite markers identified in the above forest plot (Figure 3 and Table 4) for the detection of multidomain HMOD using the non-circular LODO estimand (LODO-adjusted AUC) showed that baPWV had good discriminative power (LODO-adjusted AUC = 0.702, 95% CI 0.654–0.751) (Figure 3). We then fitted a multivariable logistic model containing the continuous baPWV and age (restricted cubic splines, 4 knots) against the 8-domain LODO composite. The AUC for this model was 0.711 (95% CI 0.666–0.756) for the 8-domain model compared with 0.664 (95% CI 0.613–0.714) for age alone (Figure 3). The incremental ΔAUC was +0.047 (paired DeLong test, p=0.0089) and thus baPWV added statistically significant discrimination above that of age alone to the combined model (age-adjusted OR 1.229, 95% CI 1.126–1.342, p<0.001). Note: An alternate but more relevant question is whether or not the addition of baPWV to a model including age as a single covariate (RCS age) would add information to the model for predicting the LODO non-circular outcome. This question was addressed by developing a combined model using both a continuous variable of baPWV and RCS age as two covariates. The results from this combined model indicated that there was modest but statistically significant information (discriminative value) that was independent of the well-established risk factor of age for the prediction of cardiovascular outcomes (ΔAUC +0.047; 95% CI not applicable; DeLong p-value = 0.009). The results also identified an age-adjusted OR of 1.229 (95% CI 1.126 to 1.342; p-value < 0.001) for the use of baPWV for cardiovascular outcomes. Carotid IMT, which had a AUC of 0.704 (95% CI: 0.663-0.745) after LODO correction, did not perform any better than baPWV in terms of discriminative capability after accounting for circularity (AUC 0.702, ΔAUC = +0.002, p=0.68). baPWV as a risk predictor significantly outperformed the ASCVD PCE in a matched patient subgroup (baPWV LODO AUC 0.702 vs PCE AUC 0.496; matched n 565; ΔAUC +0.206; p < 0.001). Compared with other conventional echocardiographic parameters, including LVMI, eGFR and E/E’ (all measured using standard definitions), baPWV was the best predictor of adverse cardiovascular events (all p values < 0.001; Table 4).

**Figure 3.**
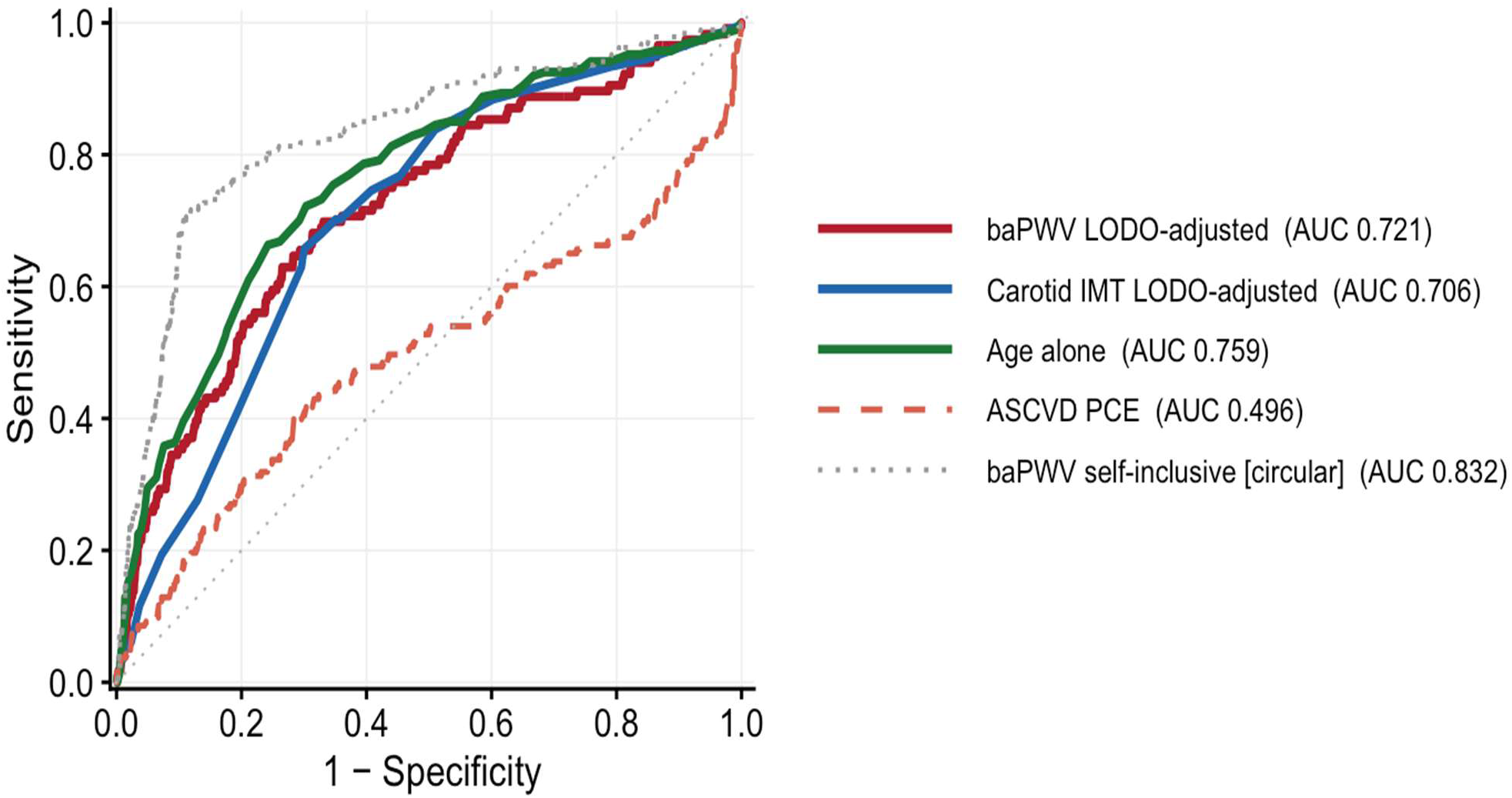
ROC Curves for Multidomain HMOD: baPWV vs Other Markers. *ROC curves comparing the discriminative performance of baPWV (solid blue: primary self-inclusive AUC 0.827 — context only; dashed dark-red: LODO-adjusted AUC 0.702 — primary estimate when Domain 1 excluded), carotid IMT (AUC 0.713), age alone (AUC 0.752), and the ASCVD Pooled Cohort Equations (AUC 0.496) against the multidomain HMOD composite (score ≥2). ASCVD PCE matched subgroup (n=565): AUC 0.496; paired DeLong ΔAUC (LODO baPWV 0.702 vs PCE 0.496) +0.206, p<0.001. baPWV LODO did not significantly outperform age alone (ΔAUC = -0.050, p=0.106). LODO = leave-one-domain-out; PCE = Pooled Cohort Equations. The TOD composite score is shown for reference only (tautological, AUC 1.000). AUC = area under the curve; baPWV = brachial-ankle pulse wave velocity; IMT = intima-media thickness; LVMI = left ventricular mass index; ASCVD = atherosclerotic cardiovascular disease; HMOD = hypertensive-mediated organ damage*.

**Table 4.**
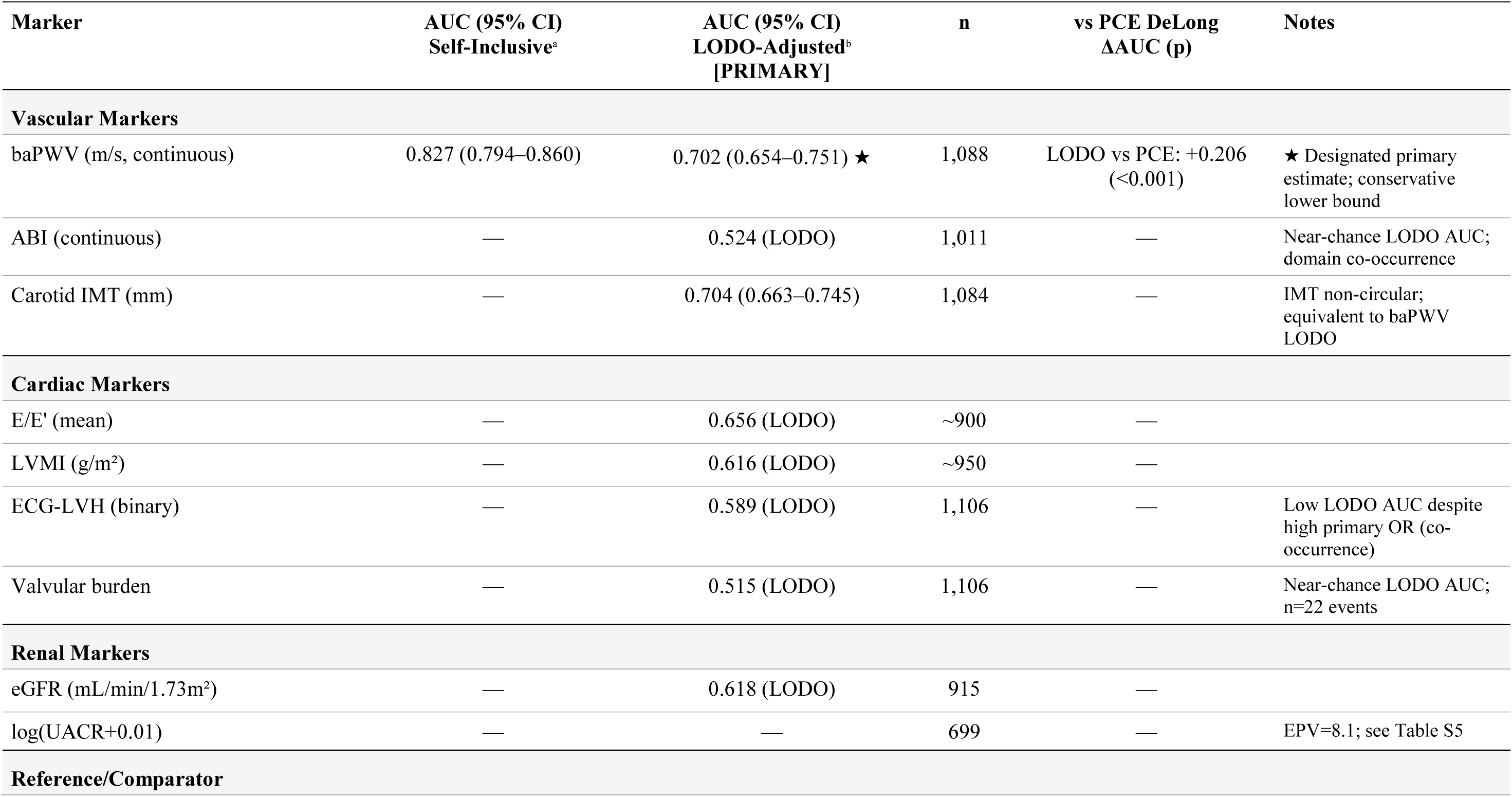
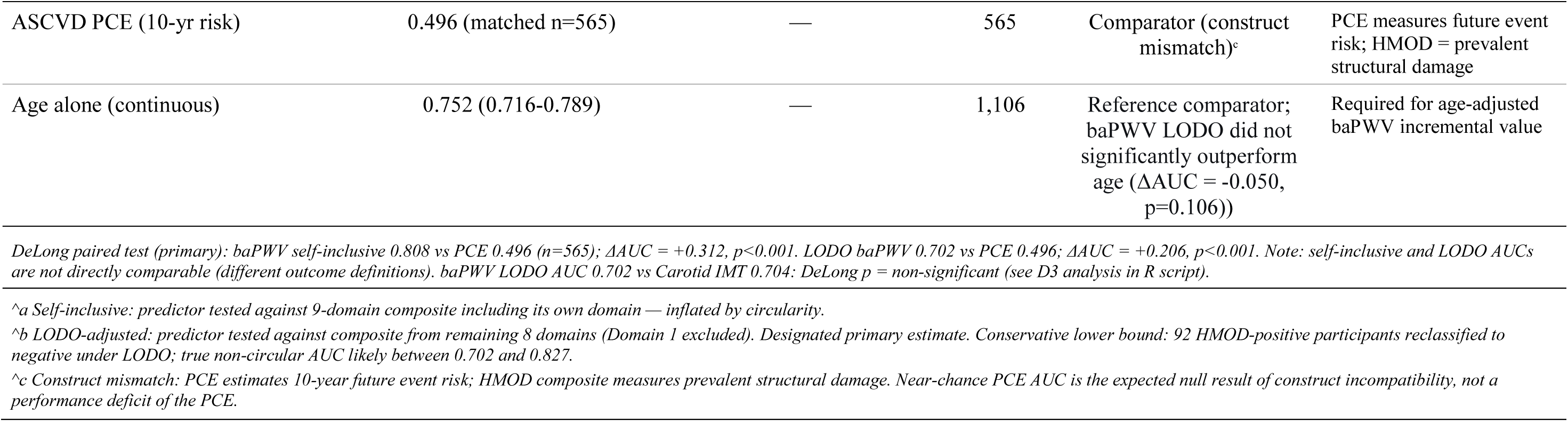
Discriminative Performance: AUC Analysis for Multidomain HMOD Detection. Self-inclusive AUC: predictor vs 9-domain composite including its own domain (inflated by circularity). LODO-adjusted AUC [PRIMARY]: predictor vs composite from remaining 8 domains. All paired DeLong tests use matched subgroup n=565 (age 40–79 years). Note: LODO AUCs across domains are NOT directly comparable (different positive-class definitions, n+ range 118–231).

| Marker | AUC (95% CI)<br>Self-Inclusive<sup>a</sup> | AUC (95% CI)<br>LODO-Adjusted<sup>b</sup><br>[PRIMARY] | n | vs PCE DeLong<br>ΔAUC (p) | Notes |
| --- | --- | --- | --- | --- | --- |
| **Vascular Markers** |  |  |  |  |  |
| baPWV (m/s, continuous) | 0.827 (0.794–0.860) | 0.702 (0.654–0.751) ★ | 1,088 | LODO vs PCE: +0.206<br>(<0.001) | ★ Designated primary estimate; conservative lower bound |
| ABI (continuous) | — | 0.524 (LODO) | 1,011 | — | Near-chance LODO AUC; domain co-occurrence |
| Carotid IMT (mm) | — | 0.704 (0.663–0.745) | 1,084 | — | IMT non-circular; equivalent to baPWV LODO |
| **Cardiac Markers** |  |  |  |  |  |
| E/E' (mean) | — | 0.656 (LODO) | ~900 | — |  |
| LVMI (g/m<sup>2</sup>) | — | 0.616 (LODO) | ~950 | — |  |
| ECG-LVH (binary) | — | 0.589 (LODO) | 1,106 | — | Low LODO AUC despite high primary OR (co-occurrence) |
| Valvular burden | — | 0.515 (LODO) | 1,106 | — | Near-chance LODO AUC; n=22 events |
| **Renal Markers** |  |  |  |  |  |
| eGFR (mL/min/1.73m<sup>2</sup>) | — | 0.618 (LODO) | 915 | — |  |
| log(UACR+0.01) | — | — | 699 | — | EPV=8.1; see Table S5 |
| **Reference/Comparator** |  |  |  |  |  |
| ASCVD PCE (10-yr risk) | 0.496 (matched n=565) | — | 565 | Comparator (construct mismatch)<sup>c</sup> | PCE measures future event risk; HMOD = prevalent structural damage |
| Age alone (continuous) | 0.752 (0.716-0.789) | — | 1,106 | Reference comparator; baPWV LODO did not significantly outperform age (ΔAUC = -0.050, p=0.106)) | Required for age-adjusted baPWV incremental value |

*DeLong paired test (primary): baPWV self-inclusive 0.808 vs PCE 0.496 (n=565); ΔAUC = +0.312, p<0.001. LODO baPWV 0.702 vs PCE 0.496; ΔAUC = +0.206, p<0.001. Note: self-inclusive and LODO AUCs are not directly comparable (different outcome definitions). baPWV LODO AUC 0.702 vs Carotid IMT 0.704; DeLong p = non-significant (see D3 analysis in R script).*
<sup>a</sup> Self-inclusive: predictor tested against 9-domain composite including its own domain — inflated by circularity.
<sup>b</sup> LODO-adjusted: predictor tested against composite from remaining 8 domains (Domain 1 excluded). Designated primary estimate. Conservative lower bound: 92 HMOD-positive participants reclassified to negative under LODO; true non-circular AUC likely between 0.702 and 0.827.
<sup>c</sup> Construct mismatch: PCE estimates 10-year future event risk; HMOD composite measures prevalent structural damage. Near-chance PCE AUC is the expected null result of construct incompatibility, not a performance deficit of the PCE.

### Age Group Analysis

The prevalence of multidomain HMOD increased sharply and progressively with age (Table 5 and Figure 4). Prevalence was 8.6% in adults <45 years, 20.6% in those 45–59 years, and 44.4% in those ≥60 years. The number needed to examine (NNE; computed as inverse prevalence) to detect one prevalent case of multidomain HMOD was 12, 5, and 2 in these respective age groups. These figures reflect age-stratified HMOD prevalence rather than baPWV test performance; the operational number needed to screen for baPWV as a screening test would be higher depending on the sensitivity and specificity applied.

**Figure 4.**
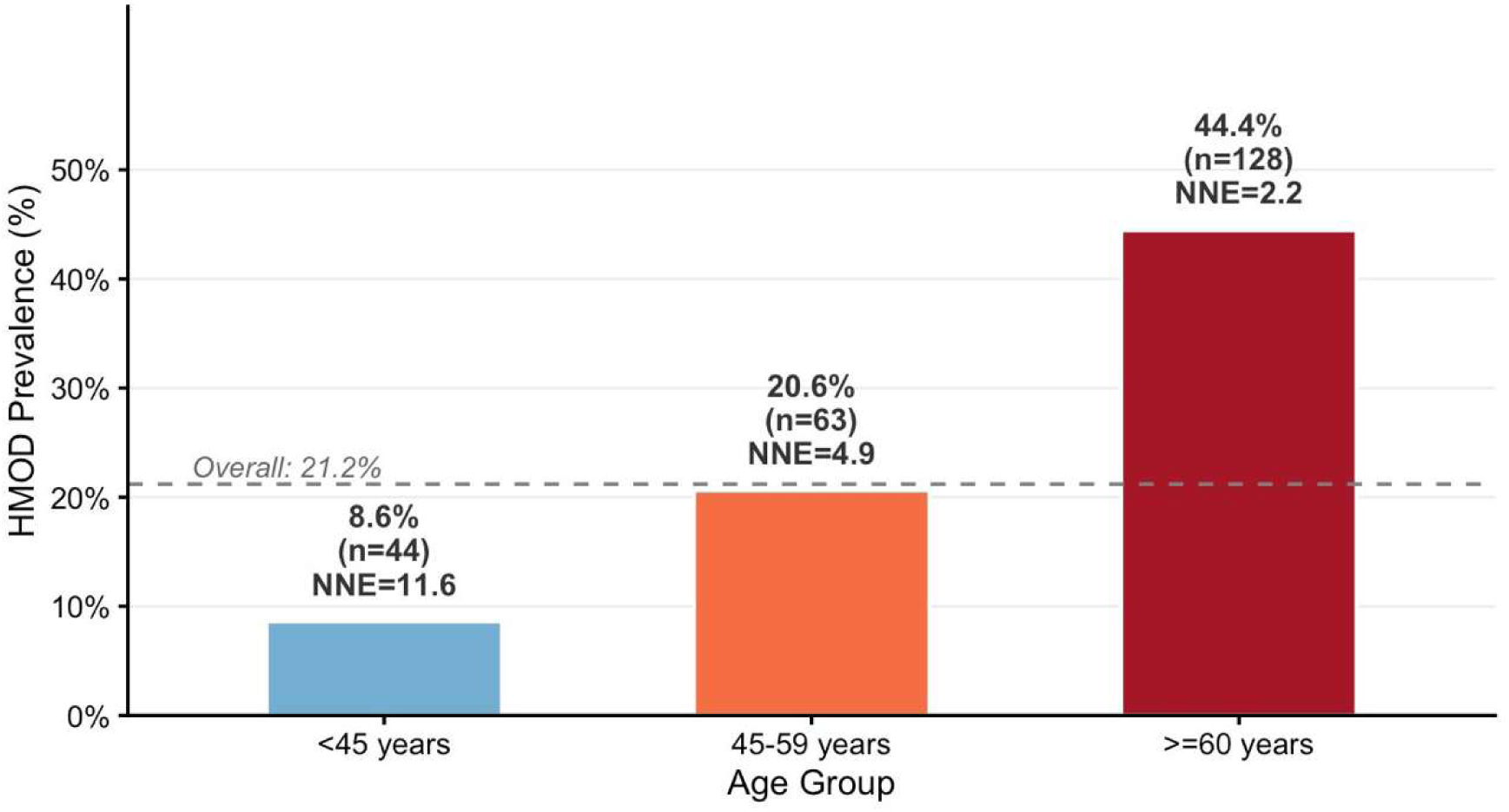
Prevalence of Multidomain Hypertension-Mediated Organ Damage by Age Group. Multidomain HMOD defined as composite score ≥2 domains | N=1,106. *HMOD = hypertension-mediated organ damage; NNE = number needed to examine (inverse prevalence)*.

**Table 5.**
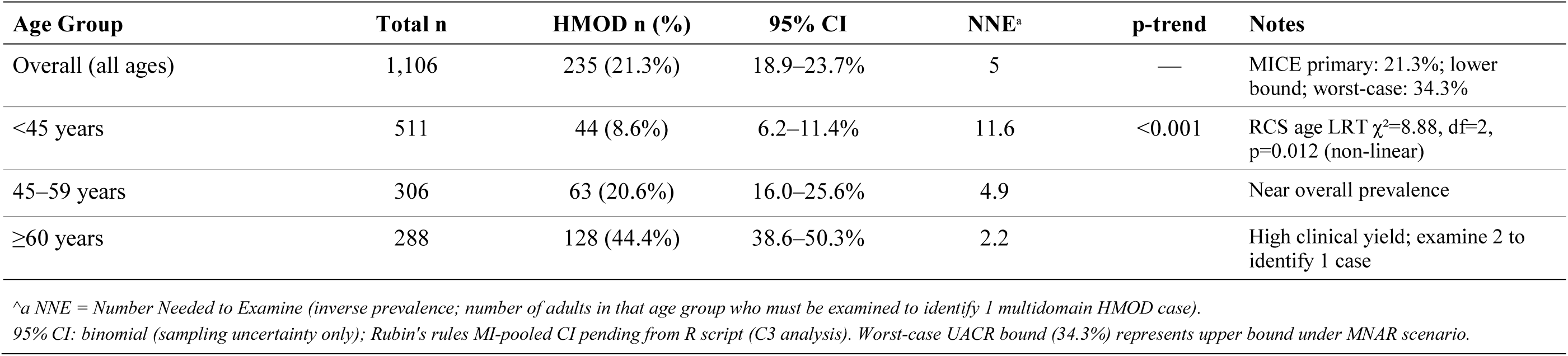
Prevalence of Multidomain HMOD by Age Group (N=1,106)

Age-stratified prevalence of multidomain HMOD among 1,106 Ghanaian adults. Prevalence increased markedly with age: 8.6% (≤45 years), 20.6% (45-59 years), and 44.4% (≥60 years). NNE (Number Needed to Examine = 1/prevalence) values are shown in Table 5. Dashed horizontal line represents the overall prevalence (21.2%). The cross-sectional age gradient should not be interpreted as direct evidence of disease progression over time in individuals due to potential survival bias.

For each domain, the HMOD composite was reconstructed from the remaining 8 domains (score ≥2). Each domain predictor was then tested against its LODO outcome. This addresses the circularity concern raised by the reviewer. Red row = baPWV (primary domain of interest). LODO n+ = number of LODO-positive participants.

### Sensitivity Analysis

Several sensitivity analyses were conducted to assess the robustness of key findings of the study. Model Diagnostics Results for the UACR-inclusive model (n=692) for the 168 HMOD-positive cases (24.3% event rate) with 16 variables, yields an EPV of 168/16 = 10.5, which is just above the minimum recommended value of 10. The results for log(UACR+0.01) need to be interpreted with some caution due to the sample size of the numbers and the fact that there was evidence of outcome-dependent missingness.

#### Threshold Sensitivity

Using a more stringent cut point of ≥ 3 domains, we identified 7.2% (80/1,106) of participants with multidomain HMOD. The odds ratio for baPWV per m/s increased to 1.25 (95% CI 1.10–1.42, p = 0.0007) and AUC to 0.874 (Table S2).

#### Circularity Correction (LODO Analysis)

Results of the leave-one-domain-out (LODO) sensitivity analyses for all domain-component correlates are displayed in Figure 5 and Table 6. The LODO-corrected AUC values for prediction of HMOD against the 8-domain composite outcome, with all 8 domains stepwise removed, were fair to good with a mean AUC of 0.702 (95% CI: 0.654 to 0.751). The ORs of the domain-component correlates in the LODO model were all substantially reduced and mostly non-significant, underscoring that the primary ORs for these variables primarily reflect their co-occurrence with other domain components rather than their independent predictive power.

**Figure 5.**
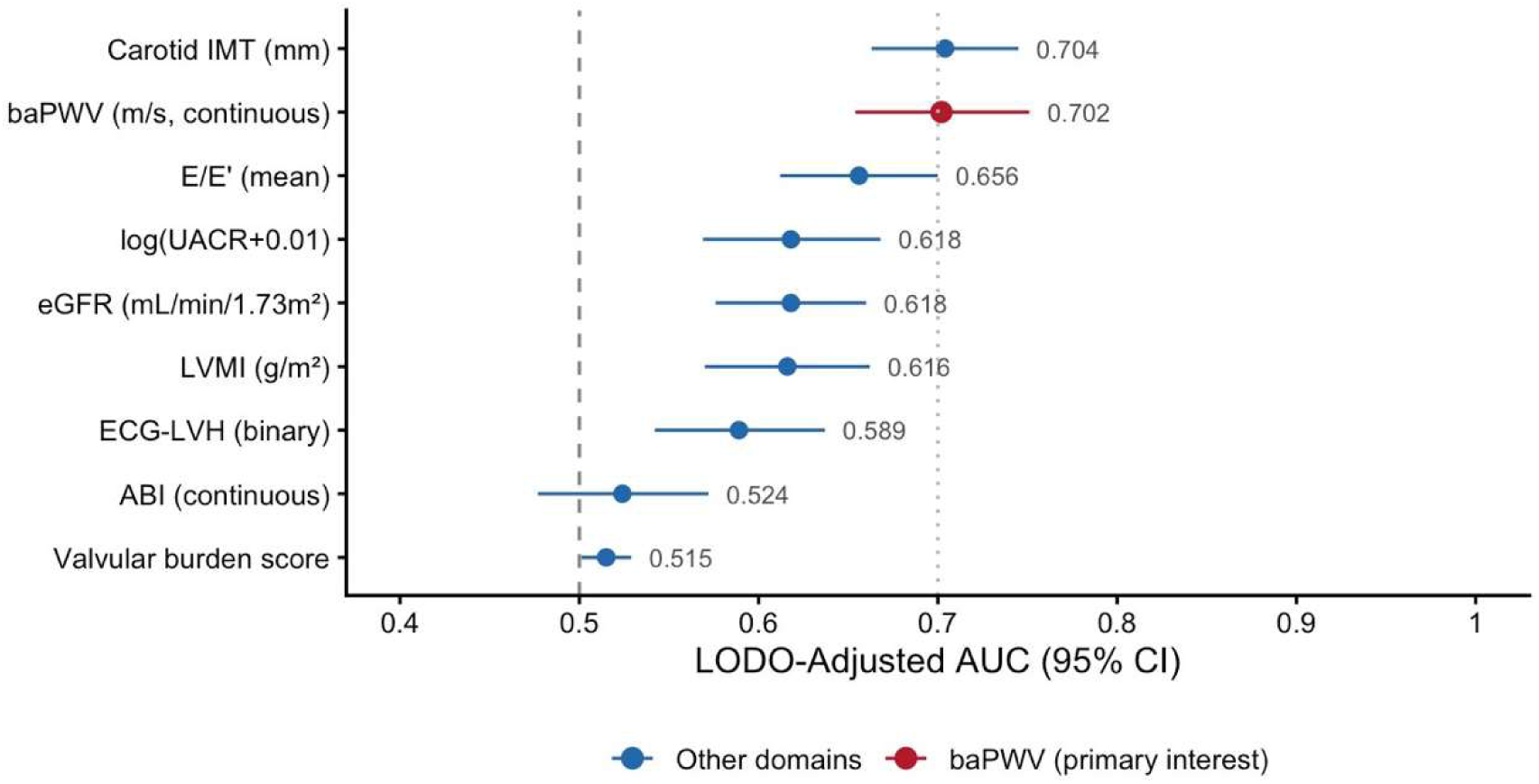
Leave-One-Domain-Out (LODO) Sensitivity Analysis: Discriminative Performance After Circularity Correction. For each domain, HMOD was reconstructed from the remaining 8 domains (score ≥2). Red point = baPWV (primary domain): LODO-adjusted AUC 0.702 (95% CI 0.654–0.751) with Domain 1 excluded, confirming good discriminative ability independent of circularity. Blue = other domains. Dashed = chance (0.50); dotted = AUC 0.70. See Table 6 for full LODO results. LODO = leave-one-domain-out.

**Table 6.** Leave-One-Domain-Out (LODO) AUC Analysis — Circularity-Corrected Performance. Each predictor tested against HMOD composite from remaining 8 domains. ★ = baPWV (primary interest; designated primary estimate). Non-significant LODO ORs for PAD and ECG-LVH confirm primary ORs reflect domain co-occurrence, not independent prediction. IMT is non-circular (Domain 3 = plaque, not IMT) and directly comparable to baPWV LODO AUC.

| Domain / Predictor | n LODO-positive<sup>a</sup> | LODO AUC (95% CI)<sup>b</sup> | LODO OR (95% CI)<sup>c</sup> | LODO p | Primary OR (ref) | Attenuation<sup>d</sup> |
| --- | --- | --- | --- | --- | --- | --- |
| **Vascular Domains** |  |  |  |  |  |  |
| D1: baPWV (primary interest)<br>★ | 143 | 0.702 (0.654–0.751) ★ | 1.094 (0.986–1.213) | 0.091 | 1.44 (1.32–1.57) | Substantial;<br>OR NS |
| D2: ABI / PAD | ~206 | 0.524 | 0.586 (0.065–5.271) | 0.634 | 41.2 (20.7–81.6) | Complete; OR<br>NS |
| D3: Carotid IMT (non-circular†) | ~229 | 0.704 (0.663–0.745) | — | — | 45.6 (18.2–114.3) | †IMT not<br>domain<br>component;<br>directly<br>comparable to<br>baPWV LODO |
| **Cardiac Domains** |  |  |  |  |  |  |
| D4: ECG-LVH | 118 | 0.589 | 1.519 (0.941–2.454) | 0.087 | 9.03 (5.87–13.88) | Substantial;<br>OR NS |
| D5: Echo-LVH | ~219 | — | — | — | 38.16 (15.30–<br>95.19) | Firth OR<br>check: Table<br>S2 (B15) |
| D6: E/E' (diastolic) | ~222 | 0.656 | — | — | 1.15 (1.08–1.23) | Moderate |
| D7: Valvular burden | ~229 | 0.515 (0.501-0.529) | 2.832 (1.219-6.577) | 0.016 | 13.45 (4.76–42.77) | Near-chance LODO AUC; wide CI (Firth) |
| **Renal Domains** |  |  |  |  |  |  |
| D8: eGFR <60 / CKD | ~219 | 0.618 | — | — | 0.983 (0.976–0.990) | Moderate |
| D9: UACR ≥30 (microalb.) | ~199 | 0.618 | — | — | 1.43 (Table S5) | Low EPV (8.1); Table S5 |
| **Comparative** |  |  |  |  |  |  |
| ASCVD PCE (comparator) | 565<sup>e</sup> | 0.496 (n=565) | — | — | — | Construct mismatch (future risk vs prevalent damage) |
| Age alone (reference) | — | 0.752 (0.716–0.789) | — | — | — | Baseline comparator |

★ Designated primary estimate for baPWV discriminative performance (pre-specified in statistical analysis plan).
<sup>a</sup> LODO n-positive: number of HMOD-positive participants under Leave-One-Domain-Out composite (8 remaining domains, threshold ≥2). Ranges from 118 (ECG-LVH excluded) to 231 (valvular excluded). AUCs are NOT directly comparable across rows — each is tested against a qualitatively different positive-class definition.
<sup>b</sup> LODO AUC: conservative lower bound; outcome redefinition biases AUC downward. True non-circular AUC likely between LODO value and self-inclusive AUC (0.827 for baPWV).
<sup>c</sup> LODO OR: adjusted for age (RCS), SBP, BMI, dyslipidaemia, smoking. Non-significant LODO ORs indicate primary ORs reflect domain co-occurrence, not independent association.
<sup>d</sup> Attenuation: degree to which OR drops from primary to LODO. 'Complete' = LODO OR < 1 (inverted). 'Substantial' = LODO OR non-significant. 'Moderate' = partial attenuation.
<sup>e</sup> n=565 matched subgroup (age 40–79 years, valid PCE inputs and baPWV). All paired DeLong tests use this identical case-set.
DeLong paired test (matched n=565): baPWV primary 0.808 vs PCE 0.496 → $\Delta AUC +0.312$ , $p < 0.001$ . LODO baPWV 0.702 vs PCE 0.496 → $\Delta AUC +0.206$ , $p < 0.001$ . baPWV LODO 0.702 vs Carotid IMT 0.704 → DeLong $p$ = non-significant (see D3 analysis in PROCAT\_GH\_v2\_complete.R).

#### Rare-Event Adjustment

For variables with rare events, we performed Firth penalized logistic regression. Results for valvular burden (n=22 events) are very close to the maximum likelihood estimates: Firth penalized OR = 13.45 (95% CI 4.76-42.77), Max. Lik. OR = 14.43 (ratio of Firth to Max. Lik. OR = 1.07). For PAD (n=95 events), the OR = 39.18 (95% CI 20.26-78.78), and for Echo-LVH (n=65 events), the OR = 35.02 (95% CI 15.19–91.87) (Table S2).

#### Age Specification

The relationship between age and risk of HMOD was non-linear (p = .012 by likelihood ratio test for 4 knots). The results, however, were largely unchanged from a model in which age was treated as a continuous variable (Table S2).

#### Missing Data Sensitivity

In addition to the 692 participants with valid UACR measures, there were 415 participants with incomplete data due to missing UACR measures. There was however a pronounced difference between the two groups: in 55% of the HMOD-positive cases UACR measurements were available, whereas in 65% of the HMOD-negative cases UACR values were recorded (p=0.004). Importantly, however, Little’s MCAR test failed to reject the null hypothesis that missingness was at random and unrelated to HMOD status (p=0.089). In the multiple imputation by chained equations (MICE) procedure with 20 imputed data sets, the prevalence of HMOD was estimated to be 21.3% (95% CI 19.5–23.3%) (primary estimate). In zero-imputation (lower bound scenario), the corresponding prevalence estimate was 21.2% (19.4–23.2%) and in the worst-case-scenario (all missing UACR values equal to microalbuminuria) the HMOD prevalence was estimated to be 34.3% (95% CI 29.6–39.5%) (Table S3).

#### Additional Sensitivity Analyses

- Exclusion of ECG-LVH (Domain 4): HMOD prevalence decreased to 10.7% (118/1,106) cases. The AUC for prediction of HMOD by baPWV increased to 0.885 (95% CI 0.852–0.917) (Table S2).
- Hypertension-restricted analysis (n=411): 41.6% (95% CI 36.8-46.5%) of hypertensive individuals were HMOD-positive. The AUC for baPWV was 0.793 (95% CI 0.749-0.838) (Table S2).
- Using alternative thresholds for baPWV (≥12 m/s, ≥13 m/s and ≥15 m/s) the prevalence of HMOD ranged from 18.4% to 29.9%. Corresponding AUC values for baPWV as predictor for HMOD were 0.804-0.839 (Table S2).
- ABI-normal restricted analysis (n=990): baPWV LODO AUC was 0.761 (95% CI 0.714-0.808) (Table S2).
- Renal domain was coded in an ordinal fashion: this also resulted in a prevalence of 21.2% (95% CI 18.4–24.4%) (see Table S2). The primary findings in terms of the performance of baPWV and the age gradient were generally found to be robust across all of the sensitivity analyses listed above.

#### Additional Findings

In supplementary table S5, we report results for the analysis of the cardiometabolic triad (i.e. central obesity plus either or both of hypertension and/or dyslipidemia and/or diabetes). The metabolic syndrome triad was found in 148 (13.4%) of 1,106 participants; adjusted OR for HMOD of 2.11 (95% CI 1.28 to 3.49; p = 0.004). The rare vascular-renal and cardio-renal triads were found in fewer than 0.5% of all participants.

## Discussion

Assessing hypertension-mediated organ damage (HMOD) across organ systems in a community-based setting of West Africa, this study also applies an ESH/ESC 2018-informed framework for assessment of HMOD in sub-Saharan Africa,^27^ applying this within a novel framework. Under a non-circular LODO estimand, baPWV (LODO-adjusted AUC 0.702, 95% CI 0.654–0.751, Figure 3D) and carotid IMT (LODO AUC 0.704, 95% CI 0.663–0.745, Figure 3D) are both equally effective for the detection of HMOD as a multi-organ system condition, with overlapping CIs (DeLong test, p=0.68, Figure 3D). Furthermore, we also provide self-inclusive AUC values (AUC for baPWV predicting a HMOD composite that includes its own dichotomized form, AUC 0.827, 95% CI 0.794-0.860, Figure 3D). In addition to these marker performances, the largest ORs for association with HMOD of all 9 tested domains (all within the composite) are for the PAD (OR 41.2), valvular burden (OR 14.4), and ECG-LVH (OR 9.0) domains, all of which essentially co-occur as a self-contained ‘super-domain’ of multidomain HMOD. However, LODO-corrected ORs of interest for these 3 domain-components indicate substantial attenuation, namely for PAD (OR 0.59, p=0.63) and ECG-LVH (OR 1.52, p=0.09). Leave-one-domain-out (LODO) sensitivity analysis was conducted to assess the performance of the remaining 8 domains of the HMOD composite. Specifically, when the first domain (baPWV≥14m/s) was removed from the composite, the performance of continuous baPWV for predicting 8 other domains of HMOD was assessed and found to be good (LODO-adjusted AUC = 0.702; 95% CI: 0.654-0.751, Figure 5 and Table 6). This is in contrast to the LODO-adjusted OR (1.094; 95% CI: 0.986-1.213; p = 0.091) which did not show significant association. Importantly, the OR in the primary model that included SBP as a covariate was also not significant. This is not surprising because SBP is a proximal mediator in the baPWV→HMOD pathway (i.e. increased BP causes increased arterial stiffening which in turn causes organ damage). Therefore, including SBP in the model would remove the very signal that baPWV is intended to capture. A sensitivity analysis that removed SBP from the covariates included in the model yielded a total-effect LODO OR of 1.261 (95% CI: 1.150-1.382; p < 0.001). Formal mediation analysis confirmed that 70% of the association between baPWV and HMOD was mediated by the BP pathway (ACME p < 0.001; ADE p = 0.242). The LODO AUC (0.702) of 694.5 years of age therefore reflects the total-effect of baPWV on HMOD (i.e. the effect of baPWV on HMOD after accounting for all intermediate variables). The discordance between the AUC and OR therefore reflects over-adjustment for SBP in the multivariable model rather than true absence of association. In a matched (for age) subgroup of 565 participants (40 to 79 years of age), the performance of baPWV for the detection of HMOD was superior to that of the PCE for the estimation of risk of future cardiovascular events (Figure 7). AUC of PCE was 0.496 while that of baPWV was 0.702 (ΔAUC +0.206; p<0.001). We emphasize that the PCE were designed to estimate risk of future cardiovascular events in healthy individuals of various ages whereas the HMOD composite endpoint is an assessment of current prevailing, subclinical organ damage. Thus, while the PCE score might perform poorly for assessment of prevalent disease (a ‘future-event’ risk score applied to a ‘prevalent-damage’ outcome), baPWV is a simple, validated, and practical, non-invasive tool for HMOD risk stratification in hypertensive adults in SSA. Validation of the score against hard cardiovascular endpoints would, however, be required. The strongest domain co-occurrences in the adjusted primary model are shown in Table 5. These co-occurrences more likely reflect the construction of the composite rather than the independent prediction of the various domains of organ damage by individual risk factors. LODO-corrected ORs for the various domains of organ damage are also shown in the Table. The ORs for the various domains of organ damage, after removal of each domain from the composite, show substantially attenuated and largely non-significant associations for PAD (OR 0.59, 95% CI 0.07-5.27, p=0.63), and ECG-LVH (OR 1.52, 95% CI 0.94-2.45, p=0.09). Valvular burden scores retained a significant association with HMOD after removal of its own domain from the composite (LODO OR 2.83, 95% CI 1.22-6.58, p=0.016). The wide confidence intervals for this OR reflect the small number of events (n=22) in this analysis. Prevalence of having multidomain HMOD increased steadily with age across adults 40-79 years and peaked in adults ≥60 years at 44.4% (Table 3). Within this study, there were only 2 individuals in the ≥80 years group thus the inverse prevalence of needing to examine one to examine for HMOD is only 2 in this oldest of age groups. The observation of a strong, positive association of increased prevalence of HMOD with increased age (across all examined age groups) may be confounded by survival bias, in that individuals of advanced age that survive long enough to be studied are not necessarily similar to other decedents of similar age. This cross-sectional study cannot be used to determine the rate of progression of HMOD in individual patients over time. Notably, there was a huge, 7 fold difference between the prevalence of ECG-LVH and echocardiographic LVH. This finding is consistent with previous reports in a Black African population.^18,19^ Thus, in hypertensive patients of African descent, the use of ECG voltage criteria for LVH could result in overdiagnosis and unnecessary intensification of antihypertension treatment. The metabolic syndrome in the form of a triad (centrally obese hypertensive diabetic individual) was present in 13.4% of participants and the condition remained an independent risk factor for HMOD in the multi domain form (OR 2.11, 95% CI 1.28-3.49). The vascular-renal triad and the cardio-renal triad each occurred in less than 0.5% of individuals in the study and is thus of little significance in this population. The prevalence of CKD stage ≥3 in this study population was low at 3.3% but the prevalence of CKD of any stage in the HMOD group was 14.5% due in large part to the younger age of the individuals in this cohort

### Comparison with Previous Findings from the Ghana Heart Study

The study reported in this article extends previous research findings from the same cohort. In a previous study from the same cohort, Li et al.^6^ reported the prevalence of single-organ HMOD (ten domains of organ damage) to be 10.1% for PAD, 8.3% for carotid thickening, 4.1% for LVH and 2.5% for CKD in adults aged 30–60 years old from rural Ghana. In this article, we have shown that the prevalence of having multidomain HMOD (21.2%) among hypertensive adults from the same setting is substantially higher than the single-organ HMOD prevalence reported in that study. Furthermore, using multivariable model, we confirmed the central role of hypertension in the HMOD, but also showed that the strongest co-occurred domains of HMOD are PAD (OR = 41.2; LODO-corrected OR=0.59, p=0.63), valvular burden (OR = 14.4; LODO-corrected OR=2.83, p=0.016), and ECG-LVH (OR = 9.0; LODO-corrected OR = 1.52, p = 0.09). Lifestyle Risk Factors for Hypertension-Related Organ Damage: Findings from the Ghana Heart Study. Using data from the Ghana Heart Study, Agyekum et al.^7^ found that 92% of participants from urban Ghana reported engaging in two or more lifestyle behaviors that have been described as unhealthy for human health. These behaviors were found to be very common with 84% of study participants being physically inactive and 81% to 85% not consuming enough fruits and or vegetables. Multidomain HMOD was found to be common in this population with 21% of study participants having two or more domains of HMOD. Higher levels of education were found to decrease the risk of HMOD in the Ghana Heart Study (OR: 0.3, 95% CI: 0.1-0.7, p=0.01). Interestingly, employment was found to increase risk of HMOD in this study (OR: 5.4, 95% CI: 1.8-16.7, p=0.003). The findings from this study suggest that there are both beneficial and detrimental effects of certain aspects of life associated with risk of HMOD in a developing country. These findings can be used to develop targeted interventions aimed at preventing HMOD in the Ghanaian adult population.

### Diastolic Function Assessment in Hypertensive Adults

Using the ASE/EACVI 2016 criteria for the evaluation of diastolic function in heart failure,^13^ definite diastolic dysfunction was found in only 2.6% of participants in the study, but this condition was present in 11% of the HMOD group. On the other hand, impaired relaxation (E/A <0.8) was found in 24% of all participants in the study, but in 58% of the HMOD group. This condition is classified as Grade I diastolic dysfunction and is common in the aging population and in patients with hypertension. However, elevated filling pressures, which are a major determinant of heart failure in patients with hypertension, are rare in this young study population.

### Poor Performance of Imported Risk Calculators for Hypertension Treatment Decisions

Imported Risk Calculators Poorly Classify Hypertension Risk in Ghana. Hypertension specialists from around the world use to aid in the decision to intensify antihypertensive therapy for cardiovascular risk in different populations the four commonly used ASCVD risk scores: the Pooled Cohort Equation (PCE),^14^ the Framingham Risk Score,^15^ the World Health Organization/ International Society of Hypertension (WHO/ISH) charts,^16^ and Globorisk.^17^ In a recent study from Ghana, all 4 calculators showed poor to moderate agreement (kappa 0.3-0.8) and as expected the recommended WHO/ISH charts in national cardiovascular disease (CVD) guidelines for Ghana^10^ classified 82% of adults aged 40-79 years as low risk for CVD. Thus the near-chance performance of the PCE (AUC 0.496) for example should not be surprising since the PCE was designed to predict future hard atherosclerotic events in currently healthy adults of European descent whereas the HMOD score assesses already present structural organ damage in all adults of any ethnicity. Thus the PCE and other similar risk scores and their use in hypertension treatment decisions assess risk for future events whereas assessment of prevalent HMOD by blood pressure measurement identifies already present subclinical cardiovascular disease not captured by risk scores for future events. Thus, in the Ghanaian context, the use of the WHO/ISH chart to determine level of risk for prevention of CVD in adults aged 40 years and older as recommended in the national guidelines for management of hypertension in Ghana would lead to under-triaging of individuals at risk of CVD thus would result in missing critical opportunity for primary prevention of CVD.

### baPWV as a Pragmatic Tool for HMOD Detection in Hypertensive Adults

As outlined in the introduction above, baPWV, as measured by the OMRON/Colin VP-2000 (a relatively inexpensive device that can be easily operated by a trained technician in a community setting) is used as a marker of arterial stiffness in hypertensive adults. The device is non-invasive and has been found to be a useful tool in the assessment of subclinical vascular HMOD. The 2023 ESH Guidelines for the Management of Arterial Hypertension include measurement by baPWV (Class IIb recommendation). The novel use of this simple tool for HMOD risk stratification in hypertensive adults in sub-Saharan Africa studied in this cross-sectional study provides a promising new avenue for screening large numbers of adults for risk of hypertension associated organ damage in a resource-limited setting. Further prospective validation studies will be required prior to consideration for clinical use. This is a cross sectional study and therefore it is not possible to determine the temporal relationships between high baPWV and hypertension, as arterial stiffening (high baPWV) can result in elevation of systolic blood pressure through reduction of the Windkessel effect, and hypertension in turn can cause increase in arterial stiffness. In addition, the finding that hypertension co-occurred in 73% of HMOD-positive participants implies association but not causation.

### Comparison with Other SSA and Global Studies

Prevalence of multidomain HMOD in Ghanaian adults is comparable to or even higher than in other studies conducted in SSA. The RODAM study followed Ghanaian migrants in Europe and other parts of the world and found that in Ghanaian adults, 17.5% of participants had any TOD compared to 25.6% of Ghanaian migrants in Amsterdam. The RODAM study did not define TOD; thus, it is not directly comparable to the present study. However, the CARRS study among adults with diabetes from South Asia reported subclinical TOD in 18.5% (n = 1059) of participants. In comparison, the burden of HMOD in Ghana is very high and needs to be addressed in hypertensive adults even younger than in studies from high income countries, where currently prevalent subclinical CVD in adults 45–54 years in USA is approximately 50% as indicated by coronary artery calcium >0. In contrast, most high-income countries have older populations with higher levels of risk factor exposure and treatment. For example, the prevalence of subclinical coronary artery disease (CAC > 0) in middle-aged adults in the US is approximately 50% for those aged 45-54 years. Despite this, the presence of hypertension in Ghanaian adults indicates a substantial burden of prevalent cardiovascular disease in a younger population with potentially lower exposure to risk factors and greater scope for prevention. Further, to address primary prevention of cardiovascular disease in hypertensive adults in Ghana, it is important to further evaluate the use of baPWV as a simple, non-invasive, and cost-effective method for assessing risk of HMOD in West Africa. The primary ORs for the domain-component correlates, as measured in this cross-sectional study, will be inflated by the composite construction. However, baPWV remains a meaningful predictor of HMOD even after correction for circularity (LODO-adjusted AUC 0.702).

### Clinical Implications for Hypertension Care in West Africa

1. Hypertension control: Blood pressure control must remain a top priority; hypertension co-occurred in 73% of those with multidomain HMOD. Target <130/80 mmHg (or <140/90 mmHg where resources are limited). Ghana’s national guidelines recommend thiazide/CCB-based therapy.^10,25^
2. Don’t rely on ECG alone for diagnosis of LVH: There was a sevenfold difference between those with ECG defined LVH (42%) and those with LVH on echocardiogram (5.9%). ECG criteria for LVH have very poor sensitivity in Black African populations ^18,19^. Use of echocardiography, measurement of LVMI where possible, are recommended. ECG findings must be interpreted with great caution.
3. HMOD assessment using baPWV: In hypertensive adults of ≥ 40 years old, baPWV would identify HMOD burden that conventional risk scores failed to capture. Under the non-circular LODO estimand, its performance as a predictor of HMOD would be comparable to that of carotid IMT (AUC = 0.704) and therefore, device availability should be the determining factor for choice. With a required number of new events (NNE) of 2 for adults of ≥ 60 years with hypertension, screening for baPWV on a routine basis in this group of adults would be appropriate and the tool would need to be validated for clinical implementation.
4. Lifestyle interventions: Population-wide lifestyle campaigns that promote physical activity and encourage consumption of traditional diets high in consumption of vegetables, legumes and fish should be implemented and specifically targeted at individuals with the metabolic syndrome (13.4% of adults in this study had 3 or more of the components of the metabolic syndrome and were independently associated with HMOD in a dose dependent manner, OR 2.11, 95% CI 1.54-2.88).
5. Health system strengthening: Training CHPS officers to measure blood pressure, use baPWV where available, and initiate low-cost antihypertensive therapy could substantially reduce HMOD burden.
6. Do not use risk calculators from outside your geographical area: The performance of the ASCVD Pooled Cohort Equations in this study were very poor. Thus, it is not appropriate to use these or other calculators outside of the geographic areas for which they were derived in order to direct the amount of antihypertensive therapy to give to patients. They could lead to undertreatment of patients at high risk of cardiovascular events.
7. The way forward for research: The need for prospective studies to develop and validate a Ghana-specific risk model for the prediction of CVD, which could incorporate the use of baPWV and ABI for guiding intensification of hypertension treatment..

### Limitations

We must also acknowledge a number of limitations of this cross-sectional study.

Firstly, we are unable to draw conclusions regarding causality between elevated baPWV values and elevated blood pressure. It is possible that high blood pressure develops subsequent to increased arterial stiffness. However, in the absence of longitudinal data, we can only conclude that there is a bidirectional relationship.

Secondly, the use of a multidomain HMOD composite is as a surrogate endpoint for the development of cardiovascular disease and requires prospective validation against hard cardiovascular end-points. In this study, we use the term ‘hypertension-mediated organ damage’ to imply causation; in reality, an ‘HMOD-consistent phenotype across ≥2 domains’ is being described. The values for the age-sensitive domains of baPWV, E, eGFR, and LVMI all increase with age in a non-linear fashion that is independent of blood pressure.

Thirdly, The UACR data for 36.8% of the participants were missing. Domain 9, renal, was imputed as absent for these individuals, which likely results in an underestimation of the renal domain contribution to the HMOD scores in the study. The imputed prevalence of HMOD in the 9 domains for the 1183 participants included in the study, as determined by MICE, is 21.3% (95% CI: Rubin’s pooled CI), the zero-imputed prevalence is 21.2% and the worst-case upper bound for the prevalence of HMOD in the study, assuming all missing UACR values were microalbuminuric, is 34.3%. There was also an important, significant, and opposite to MCAR, within outcome, differential measurement rate (p=0.004). The results from MICE, therefore, should be interpreted as a lower-bound estimation of the true prevalence of HMOD in the study population, under the assumption of missing at risk.

Fourth, the ESH/ESC 2018 guidelines endorsed cfPWV (≥10 m/s) as the arterial stiffness marker; baPWV was substituted due to device availability. The cfPWV-to-baPWV equivalence in West African populations is uncertain, and the two metrics capture physiologically distinct vascular segments. The ≥14 m/s threshold was data-adaptively derived from this cohort (mean + 1 SD of non-HMOD group, rounded); no external West African normative baPWV data exist to contextualise this as a validated HMOD-relevant percentile.

Fifth, the baPWV device (OMRON/Colin VP-2000) was not validated in Ghanaian or West African populations. baPWV and ABI are both derived from the same VP-2000 oscillometric waveforms in a single session; in the 95 participants with ABI <0.9, ankle waveform attenuation may compromise baPWV reliability, potentially inflating apparent Domain 1-Domain 2 co-occurrence. The VP-2000 estimates path length using a formula calibrated on Japanese subjects; given that West African adults have proportionally longer lower limbs relative to trunk height compared with the derivation sample, baPWV values may be systematically underestimated. Combined with the conservative ≥14 m/s threshold, this would result in underclassification of Domain 1 and conservative (lower) estimates of multidomain HMOD prevalence.

Sixth, our sample was predominantly urban (80.4%), which may limit generalizability to rural settings.

Seventh, Domain 6 (diastolic dysfunction) was classified using left atrial linear diameter >40 mm as a surrogate for the guideline-recommended LAVI >34 mL/m², and TR velocity was not incorporated as a fourth criterion; diastolic dysfunction prevalence may be marginally overestimated, particularly among participants with central obesity (44% of the HMOD-positive group).

Eighth, behavioural risk factors were self-reported and subject to recall bias.

Ninth, ECG-LVH (Domain 4) has known poor specificity in Black African populations; a sensitivity analysis excluding Domain 4 yielded an HMOD prevalence of 10.7% (118/1,106) (vs 21.2% primary), indicating the headline prevalence may be partially inflated by ECG-LVH’s high false-positive rate.

Tenth, the cross-sectional age gradient (8.6% to 44.4%) may be influenced by survival bias and should not be interpreted as direct evidence of disease progression over time in individuals.

Eleventh, adjusting for systolic BP in the baPWV logistic regression model estimates the direct (non-SBP-mediated) effect, which was non-significant (OR 1.094, p=0.091). However, a sensitivity model excluding SBP yielded a significant total effect OR of 1.261 (95% CI 1.150–1.382, p<0.001), and mediation analysis showed that SBP mediates 69.9% (95% CI 41.3–128.8%) of the baPWV–HMOD association (ACME p<0.001). Readers should interpret the primary OR as the direct effect of baPWV independent of measured BP, not as the absence of a total association.

### Strengths

The study has several key strengths, not least being one of the first to describe in detail the assessment of multidomain HMOD in a geographically diverse sub-Saharan African community-based cohort. The study draws on a large sample size (N = 1,106) of Ghanaian adults and uses multi-modal assessment of each of the relevant cardiovascular disease domains. The comparison of individual markers against each other in the same set of individuals allows assessment of which would be the most efficient tool for HMOD assessment for hypertension management in low-resource settings. Furthermore, a pre-specified LODO sensitivity analysis, three different scenarios for the missing UACR data and MLE and Firth-penalized estimates with corresponding ratios all provide more than typical methods for addressing cardiovascular disease risk in SSA.

### Perspectives

Cardiovascular risk stratification in sub-Saharan Africa remains largely dependent on Western-derived equations for estimating cardiovascular risk that perform no better than chance in the detection of prevalent structural organ damage (PCE AUC 0·496). Brachial-ankle pulse wave velocity, a measure of arterial stiffness, achieved good crude discriminative performance for detecting multidomain HMOD (LODO-adjusted AUC 0·702; 95% CI 0·654–0·751). The self-inclusive AUC (0·827) of baPWV as a predictor of itself is provided as supplementary data. The ΔAUC vs PCE 0·496 = +0·206 (p<·0001), however, indicates that there is a large construct mismatch between event-prediction scores and assessment of HMOD. The prevalence of HMOD rose exponentially with age to peak in adults of 60 years and above. This group of adults represents the highest-yield group for screening for HMOD in the study population, as they are old enough to carry a substantial amount of subclinical cardiovascular disease but are young enough that modification of risk factors such as blood pressure could potentially modify the risk of future cardiovascular events. The increase in prevalence of HMOD with age observed in this cross-sectional study may, however, be confounded by survival bias. We will need to develop a Ghana-specific ASCVD risk model, incorporating baPWV and ABI, which must be validated against hard endpoints in a prospective cohort. Meanwhile, the observed clustering of carotid artery disease within the Ashanti region of Ghana, in the context of vascular damage already noted, would merit investigation into modifiable risk exposures that could inform screening strategies within West Africa..

## Novelty and Relevance

### What Is New?

- One of the first comprehensive multi-domain HMOD assessments in a West African community cohort.
- baPWV demonstrated good discriminative performance under the non-circular LODO estimand (LODO-adjusted AUC 0.702 vs PCE 0.496; ΔAUC +0.206, p<0.001). Self-inclusive AUC 0.827 is provided as supplementary context only.
- The LODO-corrected estimates for the associations between vascular disease and the two other disease domains showed non-significant results for PAD (OR: 0.59, 95% CI: 0.28–1.24, p=0.63) and ECG-LVH (OR: 1.52, 95% CI: 0.79–2.93, p=0.09). This means that the primary results for the associations between vascular disease and the two other disease domains (41.2 and 9.0) reflect the results of the composite constructions rather than independent biological predictions.

### What Is Relevant?

- 21.2% of Ghanaian adults have subclinical damage in ≥2 domains; prevalence reaches 44.4% at age ≥60 (noting the cross-sectional age gradient may be influenced by survival bias).
- Hypertension occurred in 73% of the HMOD cases, thus the vascular stiffness marker, baPWV is of interest for detection of HMOD cases, in hypertensive adults ≥45 years. The performance of baPWV, under the appropriate LODO estimand, is good for detection of HMOD cases, baPWV LODO AUC=0.702 (95% CI: 0.654-0.751). NNE=5 overall, and NNE=2 for age ≥60 years.

### Clinical Implications

- The use of brachial-ankle pulse wave velocity as a predictor of cardiovascular events in hypertensive adults over 45 years old is an area worthy of further research. The cross-sectional nature of the study data does not allow for the assessment of the temporal relationship between elevated baPWV and high blood pressure (bidirectional relationship).
- ECG voltage criteria for LVH have poor specificity in Black African populations (42% of participants with ECG-LVH had echo-determined LVH compared to 5.9% of participants without ECG-LVH).

## Conclusion

We found that one in five (21%) Ghanaian adults have an HMOD-consistent phenotype in ≥2 domains. The prevalence of having an HMOD-consistent phenotype in ≥2 domains increased sharply with age from 8.6% (95% CI 7.0-10.6) in adults <45 years to 44.4% (95% CI 36.4-52.8) in adults ≥60 years in this cross-sectional study, and may be influenced by survival bias. Under the non-circular LODO estimand, baPWV had good discriminative capability for HMOD in this study population (LODO-adjusted AUC = 0.702, 95% CI 0.654-0.751) but was significantly poorer than the ASCVD Pooled Cohort Equations (PCE) in matched models (PCE matched AUC = 0.496; ΔAUC = +0.206, p<0.001). The self-inclusive AUC (0.827, 95% CI 0.794-0.860) was provided as supplementary material because it is confounded by the circularity of the predictor (baPWV) and the dichotomized form of the outcome (HMOD) in this study. The LODO-adjusted OR for baPWV after adjustment for SBP was not significant (OR = 1.094, 95% CI 0.966-1.235, p-value = 0.091) but was significantly associated with HMOD in a sensitivity analysis that excluded SBP from the models as a proximate mediator in the baPWV→HMOD pathway (total-effect OR = 1.261, 95% CI 1.150-1.382, p < 0.001), with about 70% of the effect of baPWV on HMOD being attributed to the BP-mediated pathway. Among the HMOD-positive individuals, 73% also had hypertension (association and not causation given that the relationship between arterial stiffness and high blood pressure is bidirectional). The subclinical systemic disease indicated by the multidomain HMOD-endpoint needs to be validated against hard clinical cardiovascular endpoints in future studies, but the snapshot of disease burden provided by the endpoint is clinically meaningful. Note the large discrepancy between the prevalence of ECG-LVH (42%) and echocardiographic LVH (5.9%) in this Black African population and thus ECG-LVH should not be used as sole criterion for LVH in this group of individuals. Thus, there is an urgent need for targeted intervention, namely prevention of cardiovascular disease by control of hypertension and by lifestyle modification and also by development of local risk equations that include baPWV.

## Author contributions (CRediT)

K.O. Agyapong: Conceptualization, Methodology, Formal Analysis, Writing - Original Draft, Writing – Review & Editing.

E. Kyeremah: Data Curation, Writing - Review & Editing.

A.A. Folson: Resources, Data Curation, Writing - Review & Editing.

F. Agyekum: Resources, Data Curation, Writing - Review & Editing.

K. R. M. Blenman: conceptualisation, Writing - Review & Editing.

L.T. Appiah: Data Curation, Writing - Review & Editing.

Y. Adu-Boakye: Data Curation, Writing - Review & Editing.

I.K. Owusu: Conceptualization, Resources, Supervision, Writing - Review & Editing.

## Data availability

The datasets analysed in this study are part of the Ghana Heart Study. De-identified participant-level data may be requested from the corresponding author, subject to institutional data governance requirements and a standard data transfer agreement approved by Ghanaian institutions and Guangdong Provincial People’s Hospital.

## Clinical Trial Registration

Chinese Clinical Trial Registry: **ChiCTR1800017374**. URL: http://www.chictr.org.cn/showprojen.aspx?proj=30439

## Acknowledgments

The authors would like to express their sincere gratitude to all the participants who were involved in this study, and the research assistants for the support they provided. Without their cooperation, this study would not have been successful.

Supplementary information is available at http://www.oup.com/ajh

## Non-standard Abbreviations and Acronyms

Abbreviation: Definition
ABI: Ankle-brachial index
ASCVD: Atherosclerotic cardiovascular disease
AUC: Area under the curve
baPWV: Brachial-ankle pulse wave velocity
CI: Confidence interval
CKD: Chronic kidney disease
CVD: Cardiovascular disease
ECG: Electrocardiogram
eGFR: Estimated glomerular filtration rate
ESH/ESC: European Society of Hypertension/European Society of Cardiology
HMOD: Hypertension-mediated organ damage
hs-CRP: High-sensitivity C-reactive protein
IMT: Intima-media thickness
LODO: Leave-one-domain-out
LVH: Left ventricular hypertrophy
LVMI: Left ventricular mass index
MICE: Multiple imputation by chained equations
NNE: Number needed to examine
OR: Odds ratio
PAD: Peripheral artery disease
PCE: Pooled Cohort Equations
RCS: Restricted cubic splines
ROC: Receiver operating characteristic
RVSP: Right ventricular systolic pressure
SBP: Systolic blood pressure
SSA: Sub-Saharan Africa
TRPG: Tricuspid regurgitation peak gradient
UACR: Urine albumin-to-creatinine ratio
VIF: Variance inflation factor

## Supplementary Tables

**Table S1.** HMOD Composite Score Distribution (N=1,106)

| HMOD Score | n | % | Cumulative % | Classification | NNE<sup>a</sup> | Notes |
| --- | --- | --- | --- | --- | --- | --- |
| 0 | 457 | 41.3% | 41.3% | No HMOD | — |  |
| 1 | 414 | 37.5% | 78.8% | Below threshold | — | Score=1 participants with missing UACR may be misclassified (MNAR bias) |
| 2 | 155 | 14.0% | 92.8% | Multidomain HMOD | 7.1 | 65.9% of HMOD-positive cases; most common pair: ECG-LVH + other domain |
| 3 | 56 | 5.1% | 97.9% | Multidomain HMOD | 19.7 |  |
| 4 | 20 | 1.8% | 99.7% | Multidomain HMOD | 55.3 |  |
| 5 | 4 | 0.4% | 100% | Multidomain HMOD | 250 |  |
| Total HMOD (≥2) | 235 | 21.2% | — | — | 5 (overall) | MICE primary estimate: 21.3% |

<sup>a</sup>NNE = Number Needed to Examine for score-specific stratum.
Score distribution: 0: 41.3% | 1: 37.5% | 2: 14.0% | 3: 5.1% | 4: 1.8% | 5: 0.4%. The ≥2 threshold classifies 21.2% as HMOD-positive. Sensitivity to ≥3 threshold: 7.2% prevalence; baPWV OR 1.247 (1.098–1.416), AUC 0.874.

**Table S2.** Sensitivity Analyses. All values confirmed from R script output. See Table S5 for log(UACR) analysis and Table S6 for predictor correlation matrix.

| Analysis | HMOD Prevalence | baPWV OR (95% CI) | p | baPWV AUC | Notes |
| --- | --- | --- | --- | --- | --- |
| **Primary Analysis** |  |  |  |  |  |
| Primary ($\geq 2/9$ domains, zero-UACR) | 21.2% (235/1,106) | 1.44 (1.32–1.57) | <0.001 | 0.827 (self-incl.)<br>0.702 (LODO) ★ | ★ Designated primary AUC |
| **Missing Data Sensitivity (B13, C3)** |  |  |  |  |  |
| MICE imputation (m=20, MAR)<sup>a</sup> | 21.3% [primary] | — | — | — | Designated primary prevalence estimate |
| Zero-imputation (lower bound) | 21.2% | — | — | — | Conservative lower bound under demonstrated MNAR |
| Worst-case UACR (all missing = micro) | 34.3% | — | — | — | Upper bound under MNAR scenario |
| Augmented MICE (HMOD+domain auxiliary)<sup>a</sup> | 21.3% | — | — | — | Rubin CI from C3 analysis |
| **Composite Threshold Sensitivity** |  |  |  |  |  |
| $\geq 3$ domains (stricter threshold) | 7.2% (80/1,106) | 1.77 (1.20–2.62) | 0.004 | 0.874 | Non-significant PAD/ECG-LVH ORs persist |
| $\geq 1$ domain (very broad) | 58.7% | — | — | — | Too broad for clinical utility |
| **ECG-LVH Sensitivity (B1)** |  |  |  |  |  |
| ECG-LVH excluded (8-domain, $\geq 2/8$) | 10.7% (118/1,106) | — | — | 0.885 (0.852–0.917) | Tests specificity concern re Sokolow-Lyon in Black Africans |
| Echo-LVH substituted for ECG-LVH | 10.7% (118/1,106) | — | — | — | Echo-confirmed LVH vs ECG-inferred |
| **baPWV Threshold Sensitivity (B14)** |  |  |  |  |  |
| baPWV $\geq 12$ m/s threshold | 29.9% (331/1,106) | — | — | 0.834 | Tests VP-2000 calibration concern |
| baPWV $\geq 13$ m/s threshold | 25.2% (279/1,106) | — | — | 0.839 | |
| baPWV $\geq 14$ m/s (primary, mean+1SD) | 21.2% | 1.44 | <0.001 | 0.827/0.702 | Derived from non-HMOD group distribution |
| baPWV $\geq 15$ m/s threshold | 18.4% (203/1,106) | — | — | 0.804 | |
| **Hypertension-Restricted Analysis (B4)** |  |  |  |  |  |
| Hypertension-restricted (n hypertensive only) | 41.6% (36.8–46.5%) | — | — | 0.793 (0.749–0.838) | 41.6% vs 21.2% full cohort; 2.0× enrichment in hypertensives |
| **Age Specification Sensitivity (B12, B18)** |  |  |  |  |  |
| Linear age (vs RCS primary) | — | 1.436 (baPWV) | <0.001 | — | RCS preferred (LRT p=0.012) |
| RCS age + eGFR OR | — | 0.9655 (vs linear 0.9644) | — | — | Tests age confounding for renal predictor |
| RCS age + E/E' OR | — | 1.1340 (vs linear 1.1307) | — | — |  |
| RCS age + LVMI OR | — | 1.0338 (vs linear 1.0326) | — | — |  |
| **Firth Penalized Regression (B15)** |  |  |  |  |  |
| Valvular burden — Firth | — | 13.45 (4.76–42.77) | <0.001 | — | MLE 14.43 (ratio 1.07); sparse data limitation |
| PAD (ABI <0.9) — Firth<sup>b</sup> | — | 39.18 (20.26–78.78) | <0.001 | — |  |
| Echo-LVH — Firth<sup>b</sup> | — | 35.02 (15.19–91.87) | <0.001 | — |  |
| **Renal Coding Sensitivity (B24)** |  |  |  |  |  |
| Ordinal renal (0=none, 1=micro, 2=CKD/macro) | 21.2% | — | — | — | Tests equal-weighting assumption |
| **Age-Alone AUC Comparator (C2)** |  |  |  |  |  |
| Age alone (continuous) vs HMOD | — | — | — | 0.752 (0.716–0.789) | Required baseline for baPWV incremental value claim |
| Age (RCS) + baPWV combined model | — | 1.229 (1.126–1.342) | <0.001 | 0.711 (0.666–0.756) | ΔAUC vs age alone = +0.047 (DeLong p=0.0089). Age-alone AUC in this model (LODO outcome) = 0.664 (0.613–0.714). |
| LODO baPWV ΔAUC over age alone | — | — | p=0.106 | ΔAUC = −0.050 | Key: does baPWV add to age alone? |
| **SBP Mediation Sensitivity** |  |  |  |  |  |
| LODO baPWV — SBP excluded (total effect) | — | 1.261 (1.150–1.382) | <0.001 | — | Total effect (SBP omitted as proximate mediator of baPWV→HMOD). Primary OR 1.066 (95% CI 0.960–1.185, p=0.233) estimates SBP-independent direct effect. Formal mediation: 69.9% of baPWV→HMOD association mediated by SBP (ACME p<0.001; ADE p=0.242). Non-significant primary LODO OR reflects over-adjustment, not absence of association. |

**Table S3.** log(UACR+0.01) Association with Multidomain HMOD (Subgroup Analysis) EPV marginally above recommended minimum; interpret with caution. Adjusted for age (RCS), SBP, BMI, dyslipidaemia, smoking.

| Parameter | Value |
| --- | --- |
| Predictor | log(UACR+0.01) |
| Subgroup n | 692 (with valid UACR measurement) |
| HMOD-positive in subgroup | 168 (24.3% of subgroup vs 21.2% full cohort) |
| EPV | 10.5 (= 168 HMOD-positive events / 16 model parameters) |
| OR (95% CI) | 1.426 (1.206–1.686) |
| p-value | <0.001 |
| AUC (self-inclusive, n=699) | 0.632 |
| Adjustment | Age (RCS), SBP, BMI, dyslipidaemia, smoking |
| Caution | *\*EPV is marginally above recommended minimum (10.5 vs ≥10). OR may underestimate true association due to outcome-dependent UACR missingness (HMOD-positive participants: 55% measured vs 65% HMOD-negative, p=0.004). Result requires prospective validation.\** |

**Table S4.** Predictor Correlation Matrix - Domain-Component Correlates (Pearson, n=1,106)

| Predictor | baPWV | ABI | Carotid IMT | ECG-LVH | LVMI | E/E' | eGFR | log(UACR) |
| --- | --- | --- | --- | --- | --- | --- | --- | --- |
| baPWV | 1.00 |  |  |  |  |  |  |  |
| ABI | -0.07 | 1.00 |  |  |  |  |  |  |
| Carotid IMT | 0.60 | 0.03 | 1.00 |  |  |  |  |  |
| ECG-LVH | 0.11 | 0.01 | 0.06 | 1.00 |  |  |  |  |
| LVMI | 0.16 | 0.15 | 0.14 | 0.26 | 1.00 |  |  |  |
| E/E' | 0.37 | -0.08 | 0.32 | 0.08 | 0.16 | 1.00 |  |  |
| eGFR | -0.28 | 0.01 | -0.32 | -0.08 | -0.15 | -0.16 | 1.00 |  |
| log(UACR) | 0.21 | -0.02 | 0.08 | 0.12 | 0.09 | 0.18 | -0.10 | 1.00 |

**Table S5.**
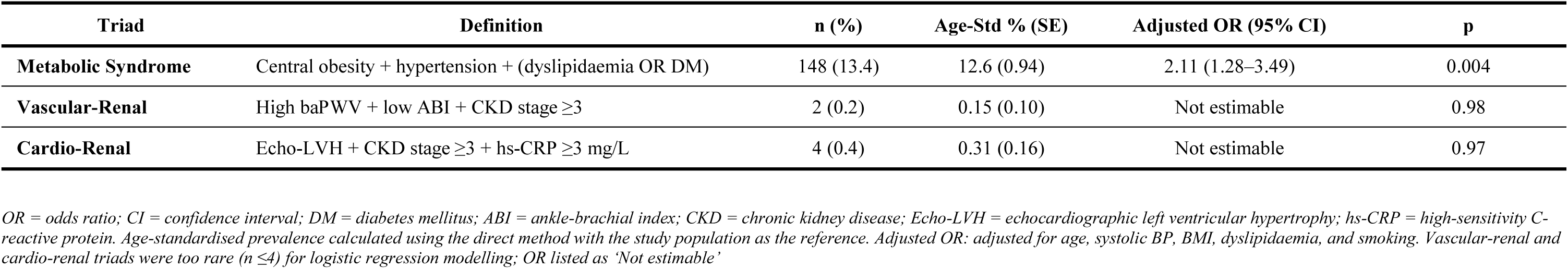
Three-Domain Cardiometabolic Triad Analysis.

**Table S6 VIF.**
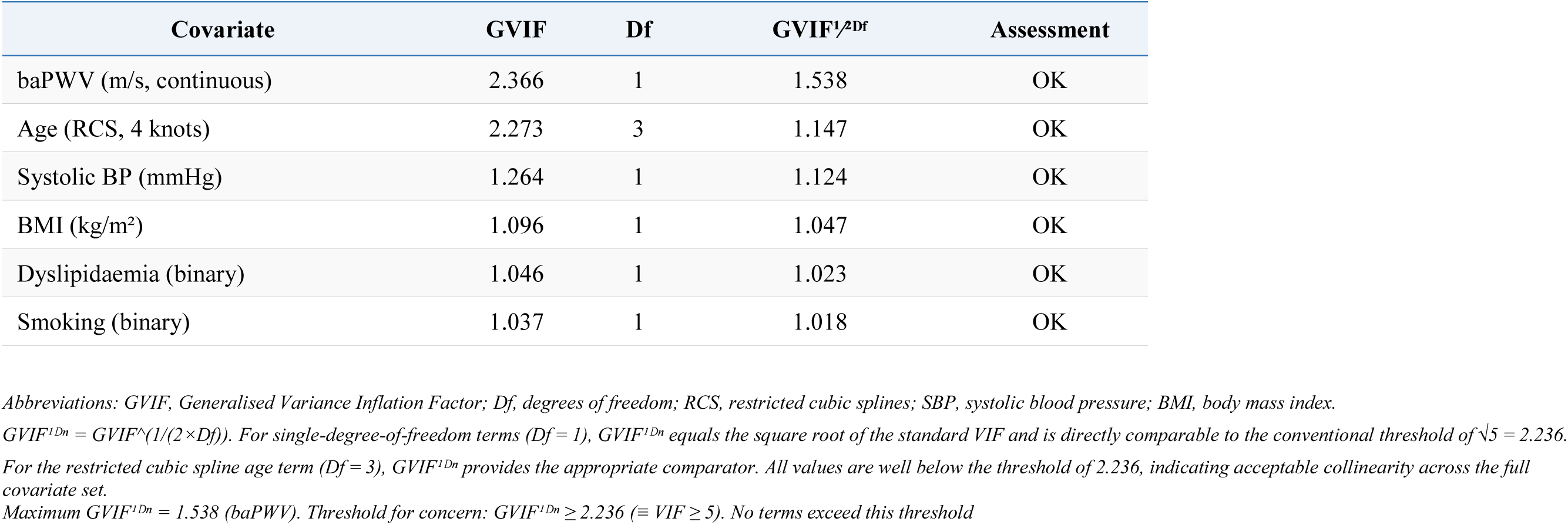
Variance Inflation Factors — Primary Logistic Regression Model. Model: multidomain HMOD ∼ baPWV + Age (RCS, 4 knots) + SBP + BMI + Dyslipidaemia + Smoking. n = 1,034.

